# Evaluation of Care Quality for Atrial Fibrillation Across Non-Interoperable Electronic Health Record Data using a Retrieval-Augmented Generation-enabled Large Language Model

**DOI:** 10.1101/2024.09.19.24313992

**Authors:** Philip Adejumo, Phyllis M. Thangaraj, Lovedeep S. Dhingra, Dhruva Biswas, Arya Aminorroaya, Sumukh Vasisht Shankar, Aline F. Pedroso, Philip M. Croon, Rohan Khera

**Author notes:** Address for correspondence: Rohan Khera, MD, MS, 195 Church St, 6^th^ Floor, New Haven, CT 06510; @rohan_khera.

## Abstract

**Importance:** Standardized assessment of clinical quality measures from electronic health records (EHRs) is challenging because information is fragmented across structured and unstructured data, and due to low interoperability across systems. Traditionally, extracting this information requires manual EHR abstraction, a time-consuming and expensive process that also limits real-time care quality improvement. **Objective:** To evaluate whether a data format-agnostic retrieval-augmented generation-enabled large language model (RAG-LLM) can accurately abstract clinical variables from heterogeneous structured and unstructured EHR data.

**Design, Setting, and Participants:** Retrospective cross-sectional study assessing stroke and bleeding risk in patients with atrial fibrillation (AF) from two health systems. We developed a RAG-LLM model to extract CHA DS-VASc and HAS-BLED risk factors from tabular data and clinical documentation.

The framework was validated on 300 expert-annotated patient records (200 from Yale New Haven Health System [YNHHS] and 100 from the Medical Information Mart for Intensive Care [MIMIC-IV]). The system was deployed on two large cohorts: 104,204 patients with AF from YNHHS (2013-2024) and 13,117 from MIMIC-IV (2008-2022). We compared anticoagulation recommendations derived from RAG-LLM with those based on traditional structured data abstraction.

**Exposures:** Use of a RAG-LLM model to abstract stroke and bleeding risk factors from structured and unstructured EHR data.

**Main Outcomes and Measures:** Accuracy of RAG-LLM-based risk factor abstraction against expert annotation. Secondary outcomes included efficiency, cross-cohort generalizability, and impact on anticoagulation eligibility based on risk stratification.

**Results:** In the validation cohort (mean age 74.8 years, 42.7% female), RAG-LLM demonstrated superior performance across all metrics compared with structural data abstraction. For individual CHA DS-VASc components, accuracy ranged from 0.94-1.00 (YNHHS) and 0.89-1.00 (MIMIC-IV) versus 0.66-0.92 (YNHHS) and 0.44-0.97 (MIMIC-IV) for structured data, which was similar for HAS-BLED (0.94-1.00 and 0.89-1.00 vs 0.66-0.94 and 0.44-0.97). In the deployment study, among 3,207 patients classified as low/intermediate stroke risk with structured data, 62.1% (1,993) were reclassified as high risk with RAG-LLM and would become eligible for anticoagulation. Similarly, 5.5% of those classified as low bleeding risk by structured data were reclassified as high risk, substantially refining contraindication assessment.

**Conclusions:** A multimodal RAG-LLM accurately abstracts clinical variables from structured and unstructured EHR data to improve stroke and bleeding risk assessments in patients with AF, enhancing identification of appropriate anticoagulation candidates.

## Background

Quality measurement in healthcare requires the comprehensive evaluation of patients’ clinical profiles from diverse data sources.^1^ Traditionally, this process has relied on manual chart review, an inefficient and expensive approach with an estimated burden of over $15 billion on the US healthcare system annually.^2–4^ Automated abstraction of structured data often misses clinically relevant information that exists within unstructured narrative documentation.^5^ Moreover, the lack of standardization and interoperability across EHR systems limits the scalability of automated quality measurement across networks.^6^ These challenges underscore the need for scalable, interoperable approaches to data abstraction for quality measurement that can capture clinical context from both structured and unstructured clinical data.

Large language models (LLMs) show promise in healthcare for interpreting clinical text and performing complex reasoning tasks.^7,8^ However, these models are prone to producing factually incorrect responses that can lead to medical errors. Additionally, their “black box” nature raises concerns about explainability and transparency in clinical applications where decision justification is critical.^9,10^ Retrieval-Augmented Generation (RAG) addresses these limitations by integrating LLMs with retrieval from vector databases, grounding outputs in source documentation to enhance transparency.^11,12^ RAG architectures excel at processing heterogeneous healthcare data by converting both structured tables and unstructured text into a unified vector space where all data elements become searchable tokens, offering a more comprehensive overview of the available data.

A widely accepted quality of care assessment in cardiovascular care is the appropriate management of anticoagulation among those with atrial fibrillation (AF). This is based on calculating CHA DS-VASc and HAS-BLED scores, which evaluate stroke and bleeding risk, respectively, to guide the use of an oral anticoagulant. These widely used risk scores require a diverse set of clinical variables as inputs, including demographics, comorbidities, laboratory values, medications, and clinical events, necessitating integration across multiple data sources.^13–15^

In this study, we developed and validated an interoperable RAG-LLM system to abstract CHA DS-VASc and HAS-BLED risk factors from structured and unstructured clinical data across two distinct US health system EHRs, using expert annotation as the reference standard. We hypothesized that this data-agnostic approach would enable fast, accurate risk assessment, outperforming approaches that use structured data alone. To assess real-world utility, we deployed the model at scale in both health systems to quantify reclassification of thrombotic and bleeding risk as a measure of anticoagulation care quality.

## Methods

The Yale Institutional Review Board reviewed the study and waived the need for informed consent due to the retrospective nature of the analysis. This study adheres to the TRIPOD (Transparent Reporting of a multivariable prediction model for Individual Prognosis or Diagnosis) reporting guidelines (**eTable 1**).^16^

### Data Sources

We used two distinct datasets from different EHR platforms to evaluate the validity and interoperability of the RAG-LLM system. The first dataset was derived from Yale New Haven Health System (YNHHS), encompassing data collected between 2013 and 2024. YNHHS operates on an Epic Clarity database architecture and includes Yale New Haven Hospital and its affiliated institutions (Bridgeport, Greenwich, Lawrence + Memorial, Saint Raphael Campus, Yale Medicine Community Hospitals, and other outpatient settings). The second dataset was obtained from the Medical Information Mart for Intensive Care (MIMIC-IV), which comprises de-identified patient records from Beth Israel Deaconess Medical Center’s emergency department and inpatient units from 2008 to 2022, and uses a Metavision EHR architecture.^17^

Both datasets included structured elements in their native tabular formats (demographics, billing codes, laboratory results, and vital signs) and unstructured elements (medical history, physical examination notes, specialty clinician notes, and discharge summaries). Each system’s native format was preserved to specifically test our framework’s ability to integrate non-standardized information (see **eFigure 1** for EHR schemas for both).

### Study Cohorts

We included all adults (≥18 years) diagnosed with AF (defined by International Classification of Diseases [ICD] 9 code 427.31 or ICD-10 codes I48.xx) and excluded patients with incomplete records. We created three distinct cohorts: development, validation, and deployment. For development, we used 100 randomly selected patients to iteratively refine our RAG queries and extraction approaches (**eFigure 2**). For validation, we randomly selected 300 patients: 200 from YNHHS (stratified across facilities to ensure representation) and 100 from MIMIC-IV and extracted their complete EHR records for expert manual annotation. Finally, to demonstrate the scalability and applicability, we deployed our RAG-LLM module on all remaining patients in both health systems.

### Study Outcomes

Our primary outcome was the accuracy of the RAG-LLM framework for abstracting clinical risk factors compared with expert-annotated reference standards. We assessed accuracy, precision (or positive predictive value), F1-score, sensitivity, specificity, and negative predictive value of the RAG-LLM model for each risk factor. Secondary outcomes included: (1) qualitative analysis of specific error types and the source of discordance between expert annotators and RAG-LLM, (2) computational efficiency comparing processing times between expert review and RAG-assisted extraction, and (3) cross-system consistency (interoperability), measuring performance variability between YNHHS and MIMIC-IV. In the deployment study, we evaluated the proportion of patients with risk factors identified by RAG-LLM versus abstraction of structured data alone, changes in risk stratification, and the proportion of patients whose anticoagulation recommendation would change based on guideline-recommended thresholds.

### Data Pre-processing

Our RAG-LLM framework employs a novel two-stage approach for automated schema mapping, transforming heterogeneous healthcare data into a unified format. This approach first automatically classifies tables into standard healthcare categories, then maps columns to standardized fields. Then, both structured and unstructured data undergo vectorization, creating a unified search space that enables the retrieval of relevant evidence across all data modalities. Technical specifications, including embedding models and processing details, are provided in the **eMethods**.

### Query Development

We developed system-agnostic queries designed to function independently of source data format through a 3-stage process. First, we extracted standardized clinical definitions from established CHA DS-VASc and HAS-BLED scoring systems and current clinical guidelines.^13,15^ Next, we manually created cross-system equivalence mappings between these clinical definitions and their corresponding structured data elements and unstructured text patterns based on expert clinical review. Finally, we integrated narrative descriptors with semantic variations to capture documentation diversity across clinical settings, incorporating feedback from iterative testing on the development cohort (**eMethods**).^13,15^ Each query incorporates both explicit structured identifiers and semantically enriched natural language patterns to accommodate terminology variations used across different EHRs (**eTables 2 & 3**).

### Model Development

Our RAG-LLM framework implements a multi-step pipeline for clinical data extraction **(Figure 1)**. The pipeline first converts diverse clinical documentation into a standardized vector space using NV-embed, a general-purpose neural embedding model, indexed in a database optimized for cosine similarity retrieval. The system then generated a structured output with supporting evidence, preserving provenance to the original documentation and storing all results in a database for full traceability. The complete technical architecture, model specifications, and retrieval parameters are detailed in **eFigure 2-3 & eTable 2-3**.

**Figure 1:**
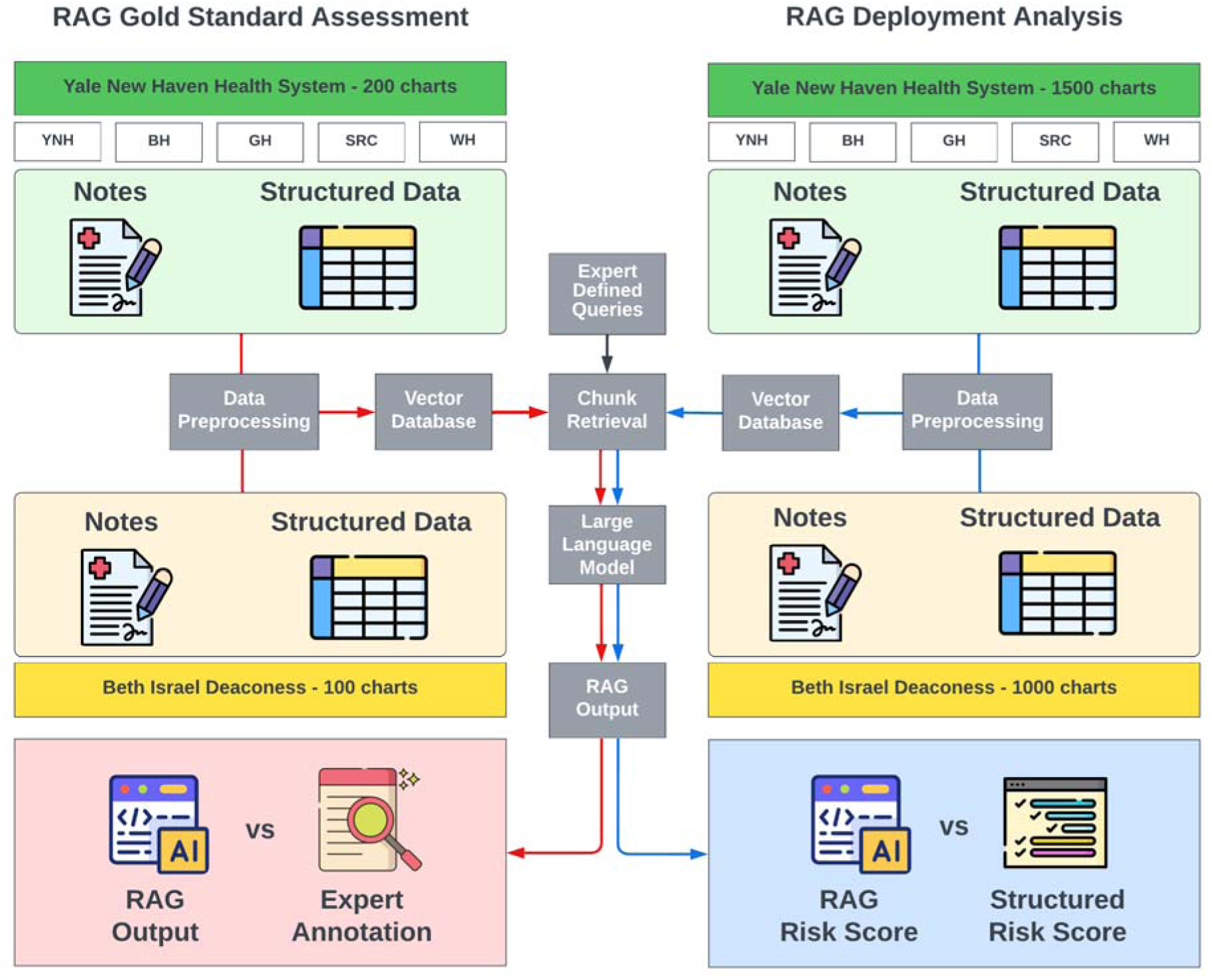
Multimodal Retrieval-Augmented Generation Framework for Integrating Structured and Unstructured EHR Data Across Non-Interoperable Health Systems. Yale New Haven Hospital (YNHH), Bridgeport Hospital (BH), Greenwich Hospital (GH), Lawrence + Memorial Hospital (LMH), Saint Raphael Campus (SRC), Westerly Hospital (WH), Medical Information Mart for Intensive Care (MIMIC-IV)

### Model Validation

In the 300-patient validation cohort, for each patient, we calculated final CHA DS-VASc and HAS-BLED scores using three distinct approaches: risk factors derived from structured data only, risk factors derived from the RAG-LLM, and the reference standard of expert review. Each record underwent an independent assessment by two expert annotators (PMT, PMC) who remained blinded to both the RAG-LLM’s outputs and each other’s determinations. This dual-annotator approach created a validation architecture specifically designed to identify potential biases or performance variations that might compromise reliability across health systems. For disagreements, a consensus panel comprising the two expert annotators and all clinician co-authors (RK, DB, LD, and AA) established the final group determination for each discordant label, forming a consensus reference standard. Error patterns of both human annotators and RAG-LLM were recorded. Processing efficiency was measured via annotator-reported time per batch (using stopwatches) and automated logs capturing RAG-LLM query durations, with total time including all steps from data retrieval to result storage. Each patient required assessment of 12 distinct risk factors (6 CHA DS-VASc and 6 HAS-BLED components), resulting in 2,400 determinations for YNHHS and 1,200 for MIMIC-IV. Performance metrics were calculated for each risk factor against the consensus reference standard. Detailed annotation protocols, adjudication procedures, and error categorization methodology are provided in **eTables 8 & 9**.

### Calculation of Risk Scores and Assessment of Anticoagulation Therapy

In the full deployment cohorts, we calculated CHA DS-VASc and HAS-BLED scores using two approaches: risk factors derived from structured data abstraction and risk factors derived from the RAG-LLM. We categorized CHA DS-VASc scores as low-risk (score 0), intermediate-risk (score 1), or high-risk (score ≥2 in men, ≥3 in women) and HAS-BLED scores as low-risk (0-1), intermediate-risk (2), or high-risk (≥3), according to current guidelines.^18,19^ We then assessed the proportion of patients whose risk categories changed when using RAG-LLM compared with structured data alone, and evaluated the implications for anticoagulation recommendations. We classified patients as not receiving oral anticoagulation at the index AF encounter if there was no active inpatient administration of warfarin or a direct-acting oral anticoagulant and no outpatient prescription supply overlapping the index date. Patients on prophylactic-dose heparins or peri-procedural, time-limited anticoagulation were not considered to be on anticoagulation therapy.

## Statistical Analysis

Descriptive statistics summarized baseline characteristics using counts and percentages for categorical variables and mean and standard deviation, or median and interquartile range for continuous variables. Accuracy of both the RAG-LLM approach and the structured data-only extraction method against the expert review was assessed by the sensitivity, specificity, positive predictive value, negative predictive value, and overall accuracy for each component of the CHA DS-VASc and HAS-BLED scores. For the deployment analysis, we examined proportions of patients shifting between risk categories (low, intermediate, high) for both stroke and bleeding risk when comparing structured versus the RAG-LLM approach. For all outcomes, we calculated 95% confidence intervals using bootstrap resampling with 1,000 iterations for population-level estimates, and the Wilson score method for individual proportions to account for smaller sample sizes in subgroup analyses. All analyses were performed using Python version 3.9, and statistical significance was set at 0.05 for all comparisons.

## Results

### Cohort Characteristics

In the YNHHS cohort (n=104,204), mean age was 76.2 years (SD±13.2) with 43.1% female. Patients were predominantly White (85.4%), followed by Black (6.7%), Asian (1.0%), and other or unknown race (14.1%). In the MIMIC-IV cohort (n=13,117, mean age 72.3±12.4 years, 42.3% female), the racial distribution consisted of 69.7% White, 7.7% Black, 2.5% Asian, and 20.1% of other or unknown race (**Table 1**).

**Table 1.** Patient Demographics and Comorbidities by Health System Cohort. Comparison of demographic characteristics and prevalence of key comorbidities across Yale New Haven Health System (YNHHS) facilities (YNH=Yale New Haven Hospital, BH=Bridgeport Hospital, GH=Greenwich Hospital, LMH=Lawrence + Memorial Hospital, SRC=Saint Raphael Campus, WH=Westerly Hospital) and Medical Information Mart for Intensive Care (MIMIC-IV) database.

| Characteristic | Overall YNHHS | YNH | BH | GH | LMH | SRC | YM-Community | Other | MIMIC-IV |
| --- | --- | --- | --- | --- | --- | --- | --- | --- | --- |
| Number of patients | 104,204 | 42,811 | 19,951 | 11,260 | 9,401 | 4,724 | 6,814 | 9,243 | 13,117 |
| Age, mean ± SD (years) | 76.2 ± 13.2 | 75.2 ± 13.0 | 76.0 ± 12.9 | 78.3 ± 12.7 | 75.7 ± 12.8 | 76.5 ± 13.9 | 75.6 ± 12.8 | 79.0 ± 14.7 | 72.3 ± 12.4 |
| Female sex | 44,948 (43.1%) | 18,001 | 8,755 | 4,982 | 3,966 | 2,295 | 2,964 | 3,985 | 5,548 (42.3%) |
| Race/Ethnicity |  |  |  |  |  |  |  |  |  |
| White/Caucasian | 89,005 (85.4%) | 36,819 | 16,046 | 10,074 | 8,474 | 3,578 | 5,919 | 8,095 | 9,141 (69.7%) |
| Black/African American | 6,954 (6.7%) | 3,233 | 1,663 | 287 | 371 | 846 | 298 | 256 | 1,014 (7.7%) |
| Asian | 1,018 (1.0%) | 389 | 145 | 178 | 103 | 38 | 76 | 89 | 322 (2.5%) |
| Other/Unknown | 14,676 (14.1%) | 5,802 | 3,870 | 1,043 | 870 | 1,144 | 852 | 1,095 | 2,640 (20.1%) |
| Comorbidities |  |  |  |  |  |  |  |  |  |
| Hypertension | 86,506 (83.0%) | 35,446 | 16,602 | 9,343 | 7,780 | 3,929 | 5,695 | 7,711 | 10,965 (83.6%) |
| Diabetes | 32,611 (31.3%) | 13,373 | 6,280 | 3,560 | 2,986 | 1,455 | 2,130 | 2,827 | 4,581 (34.9%) |
| Heart Failure | 45,330 (43.5%) | 18,559 | 8,705 | 4,958 | 4,102 | 2,069 | 2,909 | 4,028 | 6,443 (49.1%) |
| Stroke/TIA | 23,309 (22.4%) | 9,575 | 4,391 | 2,506 | 2,111 | 1,069 | 1,525 | 2,132 | 1,632 (12.4%) |
| Vascular Disease | 52,917 (50.8%) | 21,780 | 10,103 | 5,623 | 4,756 | 2,352 | 3,483 | 4,820 | 3,383 (25.8%) |
| Renal Disease | 17,048 (16.4%) | 7,004 | 3,264 | 1,842 | 1,538 | 773 | 1,115 | 1,512 | 4,407 (33.6%) |
| Liver Disease | 6,441 (6.2%) | 2,646 | 1,233 | 696 | 581 | 293 | 421 | 571 | 826 (6.3%) |
| Bleeding History | 20,611 (19.8%) | 8,469 | 3,946 | 2,203 | 1,841 | 932 | 1,342 | 1,878 | 944 (7.2%) |
| Alcohol Abuse | 13,142 (12.6%) | 5,401 | 2,515 | 1,404 | 1,186 | 596 | 859 | 1,181 | 778 (5.9%) |
| Labile INR | 3,248 (3.1%) | 1,334 | 622 | 352 | 293 | 147 | 212 | 288 | 231 (1.8%) |

### Risk Factor Identification in the Validation Cohort

Manual review of the 300-patient validation cohort confirmed a high prevalence of risk factors (**eTable 10**). Structured data extraction alone identified only a subset of clinically documented risk factors, with variable sensitivity across components. In YNHHS, structured data identified 83.9% (73/87) and 79.5% (31/39) of individuals with hypertension and diabetes, respectively. Moreover, sensitivity for vascular disease was 83.3% (45/54), and for the detection of prior stroke or TIA was 50.0% (13/26). In MIMIC-IV, structured data identified 95.3% (82/86) of patients with hypertension, 100.0% (52/52) of those with diabetes, and 100.0% (66/66) of patients with vascular disease (**eTable 10**).

The RAG-LLM demonstrated consistently higher sensitivity across both cohorts. In YNHHS, the RAG-LLM identified 97.9% (143/146) of those with hypertension, 98.3% (116/118) of vascular disease, and 96.8% (92/95) of diabetes. The RAG-LLM was also able to identify 89.3% (25/28) of prior stroke/TIA cases, representing a 78.6% higher identification over structured data alone. For bleeding risk assessment, RAG-LLM identified 95.9% (71/74) of those with renal disease, and 100% (24/24) of those with labile INR. Similarly, in MIMIC-IV, the RAG-LLM identified 97.7% (84/86) of those with hypertension, 98.1% (51/52) with diabetes, and 98.1% (51/52) with vascular disease.

For CHA DS-VASc components, RAG-LLM accuracy ranged from 0.94-1.00 in YNHHS and 0.89-1.00 in MIMIC-IV, compared with 0.66-0.92 for methods using only structured data extraction in YNHHS and 0.51-0.97 in MIMIC-IV (**eTable 5**). The system maintained high precision (0.78-1.00) while achieving sensitivity of 0.87-1.00 across all risk factors. The most pronounced difference in sensitivity was in vascular disease (0.83 vs. 0.98 in YNHHS) and identification of prior stroke/TIA (0.50 vs. 0.89 in YNHHS & 0.84 vs. 0.89 in MIMIC-IV). Compared with structured abstraction, the sensitivity of RAG-LLM was similarly higher for the HAS-BLED components. The most notable differences were in the identification of labile INR (0.17 vs 1.00 in YNHHS and 0.22 vs 1.00 in MIMIC-IV), liver disease (0.08 to 0.97), and alcohol use (0.08 to 0.92). For HAS-BLED components, RAG-LLM accuracy ranged from 0.94-1.00 in YNHHS and 0.89-1.00 in MIMIC-IV versus 0.66-0.94 (YNHHS) and 0.44-0.97 (MIMIC-IV) for structured data. The RAG-LLM approach consistently had higher or equivalent sensitivity compared with structured abstraction across all risk factors (**Figure 2 & eTable 6**).

**Figure 2.**
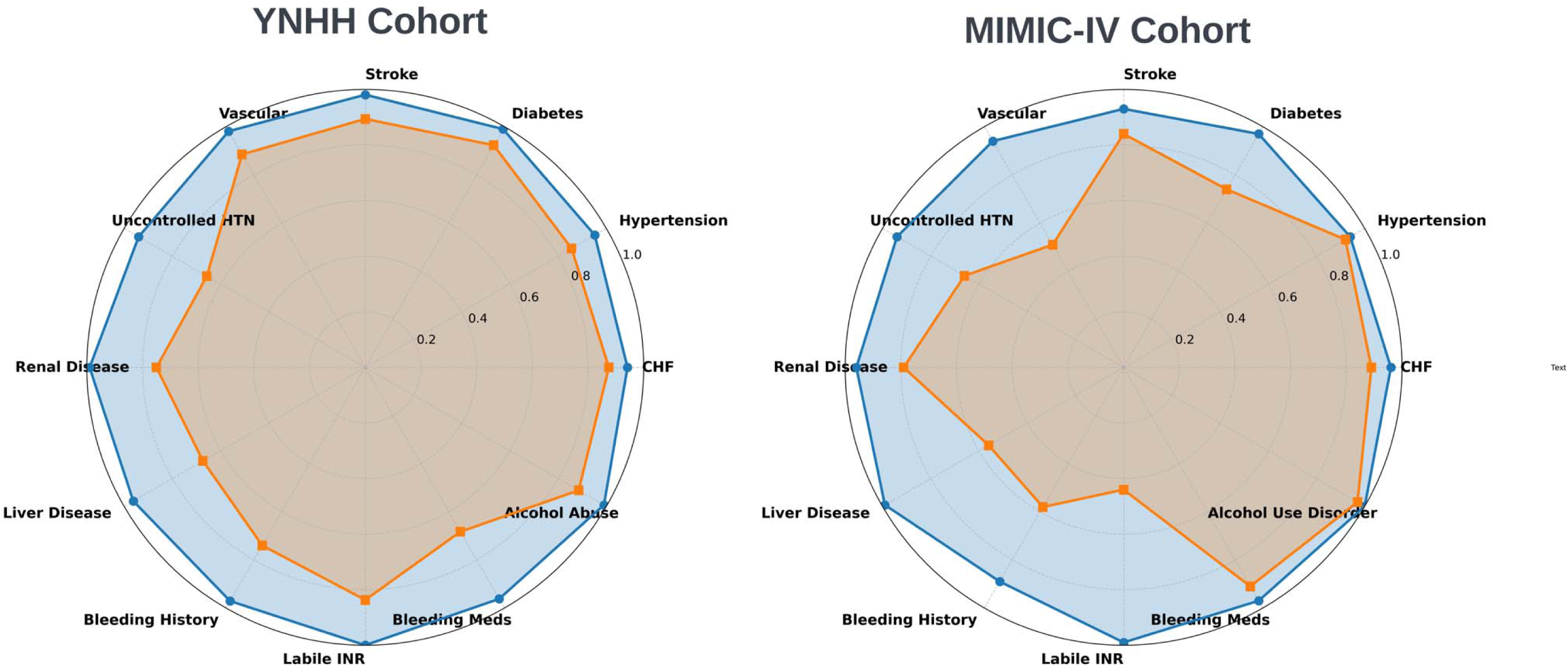
Accuracy Comparison of Retrieval-Augmented Generation (RAG) versus Structured Data Extraction for Risk Factor Identification. Radar plot illustrating accuracy for components of CHA DS-VASc and HAS-BLED risk assessment tools across both cohorts.

### Abstraction Efficiency

The RAG-LLM approach markedly outperformed expert chart abstraction in efficiency. Expert annotation required an average of 12 minutes per patient for complete assessment of all 12 risk factors, totaling 60 person-hours to review the 300-patient validation cohort. In contrast, RAG-LLM processed each patient in approximately 35 seconds, completing the full cohort in 2.9 hours, a 21-fold improvement in processing speed (**eTable 7**).

### Qualitative Assessment of the Pattern of Expert vs RAG-LLM Errors

In the YNHHS validation cohort, a comprehensive assessment of 2,400 risk factors revealed high concordance between RAG-LLM and expert annotation, with agreement in 2,196 (91.5%) instances. Among the 204 (8.5%) discordant assessments, expert adjudication determined that RAG-LLM was correct in 78 (38.2%) cases, representing instances where expert annotation had erroneously classified the risk factor. Similarly, in the MIMIC-IV cohort, assessment of 1,200 risk factors showed concordance in 1,073 (89.4%) instances. Among the 127 (10.6%) discordant assessments, RAG-LLM was correct in 42 (33.1%) (**eTable 8**).

Specific patterns of error were identified in both individual manual annotations and RAG-LLM assessments, compared with the final panel abstraction. For expert annotation, reviewers most frequently missed laboratory values marginally exceeding diagnostic thresholds (41.0% in YNHHS, 40.5% in MIMIC), explicit disease classifications documented in subspecialty notes (28.2% in YNHHS, 28.6% in MIMIC), and complex vascular conditions requiring subspecialty interpretation (19.2% in YNHHS, 19.0% in MIMIC). For RAG-LLM, the most common error patterns included contextual misinterpretation of isolated clinical findings (27.8% in YNHHS, 28.2% in MIMIC), challenges with temporal disambiguation (22.2% in YNHHS, 22.4% in MIMIC), and challenges distinguishing between acute and chronic conditions (19.0% in YNHHS, 18.8% in MIMIC) (**eTable 9**).

### Risk Factor Identification in Large-Scale Deployment

When deployed across the full YNHHS cohort (N=104,204), the RAG-LLM identified higher prevalences of CHA DS-VASc and HAS-BLED risk factors compared with structured data alone (**Table 2**). For CHA DS-VASc components, the largest difference was in detecting stroke/TIA history, where RAG-LLM identified 35,560 patients compared with 23,309 using structured data, a 52.6% relative difference. Similar patterns were observed for vascular disease (54,003 vs 52,917 patients), heart failure (52,652 vs 45,330 patients), and diabetes (34,361 vs 32,611), while hypertension showed minimal difference (87,516 vs 86,506).

**Table 2.** Differential Risk Factor Identification Between Retrieval-Augmented Generation (RAG) and Structured Data Approaches in the Deployment Subset.

|  | YNHHS |  |  | MIMIC-IV |  |  |
| --- | --- | --- | --- | --- | --- | --- |
| **Risk Factor Component** | **Structured Data** | **RAG-LLM** | **Relative Increase** | **Structured Data** | **RAG-LLM** | **Relative Increase** |
| **CHA<sub>2</sub>DS<sub>2</sub>-VASc Components** |  |  |  |  |  |  |
| **Congestive Heart Failure** | 45,330 (43.5%) | 52,652 (50.5%) | **+16.2%** | 6,443 (49.1%) | 6,551 (49.9%) | **+1.7%** |
| **Hypertension (CHA<sub>2</sub>DS<sub>2</sub>-VASc)** | 86,506 (83.0%) | 87,516 (84.0%) | **+1.2%** | 10,965 (83.6%) | 11,112 (84.7%) | **+1.3%** |
| **Diabetes** | 32,611 (31.3%) | 34,361 (33.0%) | **+5.4%** | 4,581 (34.9%) | 5,248 (40.0%) | **+14.6%** |
| **Stroke/TIA** | 23,309 (22.4%) | 35,560 (34.1%) | **+52.6%** | 1,632 (12.4%) | 3,557 (27.1%) | **+118.0%** |
| **Vascular Disease** | 52,917 (50.8%) | 54,003 (51.8%) | **+2.1%** | 3,383 (25.8%) | 7,660 (58.4%) | **+126.4%** |
| **HAS-BLED Components** |  |  |  |  |  |  |
| **Uncontrolled Hypertension** | 12,823 (12.3%) | 13,597 (13.0%) | **+6.0%** | 1,658 (12.6%) | 3,247 (24.8%) | **+95.8%** |
| **Renal Disease** | 17,048 (16.4%) | 22,403 (21.5%) | **+31.4%** | 4,407 (33.6%) | 6,931 (52.8%) | **+57.3%** |
| **Liver Disease** | 6,441 (6.2%) | 9,812 (9.4%) | **+52.3%** | 826 (6.3%) | 2,047 (15.6%) | **+147.8%** |
| **Bleeding History** | 20,611 (19.8%) | 24,628 (23.6%) | **+19.5%** | 944 (7.2%) | 3,498 (26.7%) | **+270.6%** |
| **Bleeding Medications** | 18,512 (17.8%) | 21,701 (20.8%) | **+17.2%** | 6,211 (47.4%) | 9,738 (74.2%) | **+56.8%** |
| **Labile INR** | 3,248 (3.1%) | 4,789 (4.6%) | **+47.4%** | 231 (1.8%) | 421 (3.2%) | **+82.3%** |
| **Alcohol Use** | 13,142 (12.6%) | 14,127 (13.6%) | **+7.5%** | 778 (5.9%) | 904 (6.9%) | **+16.2%** |

For HAS-BLED components, differences with RAG-LLM were observed across multiple domains, with 21,701 patients on bleeding-predisposing medications versus 18,512 by structured data (17.2% higher). Uncontrolled hypertension was detected in 13,597 patients versus 12,823 (6.0% higher), and bleeding history in 24,628 versus 20,611 (19.5% higher). Additional differences were seen for liver disease (9,812 vs. 6,441; 52.3% higher), labile INR (4,789 vs 3,248; 47.4% higher), and renal disease (22,403 vs. 17,048; 31.4% higher). Alcohol use showed minimal difference, with 14,127 patients identified by RAG-LLM versus 13,142 with structured data alone.

These results were consistent in the MIMIC-IV deployment cohort (N=13,117), confirming the generalizability of RAG-LLM-based risk factor identification (**eTable 10**).

### Changes in Risk Stratification Between RAG-LLM and Structured Data

The RAG-LLM approach resulted in substantial shifts in both stroke and bleeding risk stratification in the full YNHHS cohort. For CHA DS-VASc-based stroke risk assessment, 2,290 (2.2%) of patients changed risk categories based on the RAG-LLM data extraction **(eTable 11)**. Among patients initially classified as low or intermediate stroke risk by structured data (n=3,207), nearly two-thirds were reclassified to high risk with RAG-LLM (1,993/3,207; 62.1%). Overall, 1,993 patients (1.9%) moved from low/intermediate to high risk, while 184 (0.2%) moved from high to lower risk categories.

For HAS-BLED bleeding risk assessment, 36,866 (35.4%) patients were reclassified to a different risk category. Among patients initially categorized as low risk (n=50,774), 16,707 (33.0%) were reclassified to higher risk levels, with 13,901 (27.4%) to intermediate risk and 2,806 (5.5%) to high risk. In the intermediate-risk group (n=32,406), 4,741 (14.6%) were reclassified to low risk and 11,764 (36.3%) to high risk. Among those initially classified as high-risk (n=21,024), 3,654 (17.4%) were reclassified to a lower risk category **(Figure 3, eTable 12 & eFigure 4)**.

**Figure 3.**
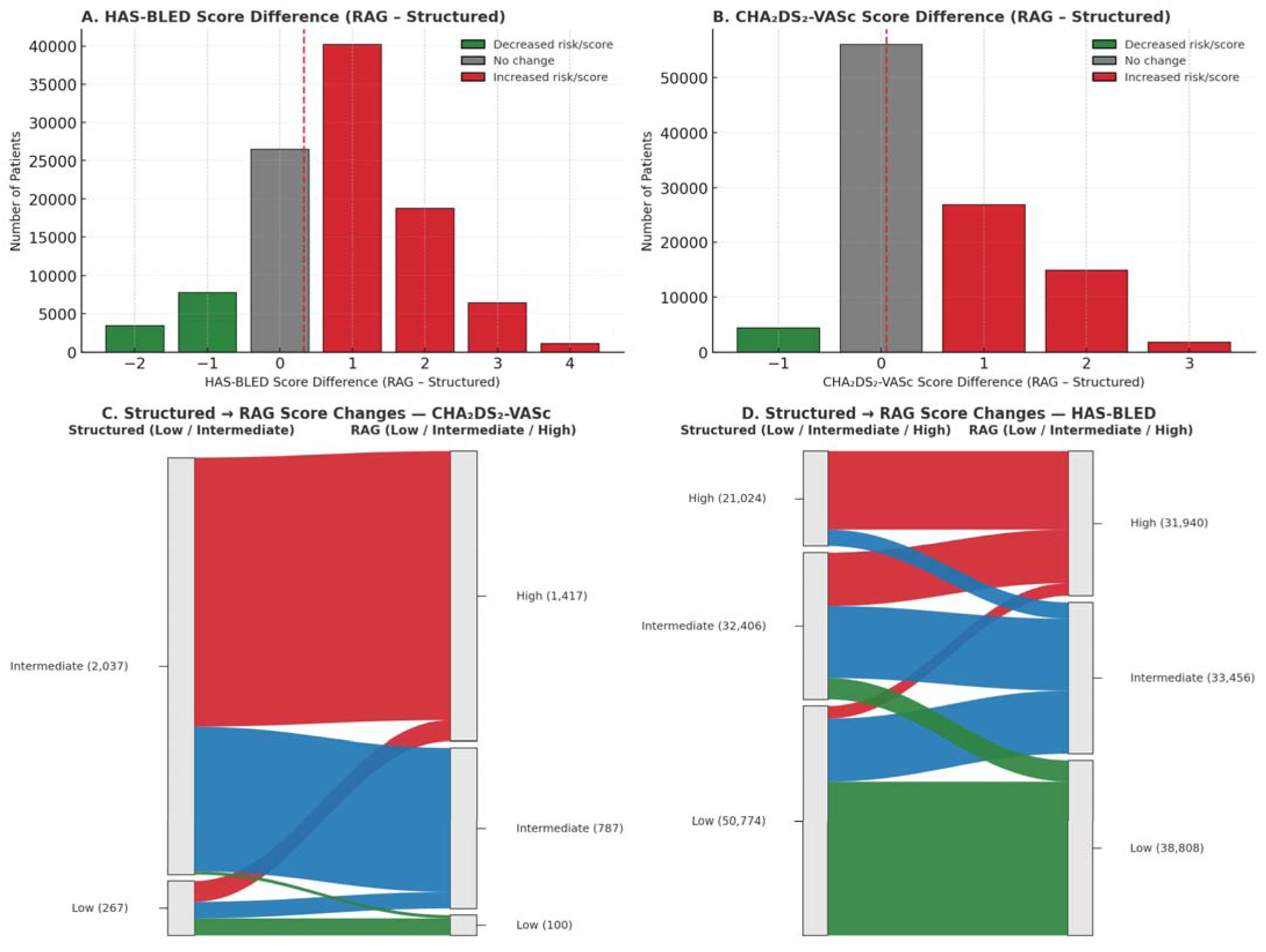
**Sankey Diagram Showing Patient Transitions Between Risk Categories for Stroke Risk (CHA DS-VASc) and Bleeding Risk (HAS-BLED) After Implementation of Retrieval-Augmented Generation (RAG).** (A) HAS-BLED and (B) CHALJDSLJ-VASc score differences (RAG − Structured) in the YNHHS deployment cohort. (C–D) NO OAC subset transitions from Structured Low/Intermediate to RAG Low/Intermediate/High for (C) CHALJDSLJ-VASc and (D) HAS-BLED

These changes would substantially alter treatment recommendations. Among individuals for whom structured data abstraction would not prompt anticoagulation because of low/intermediate stroke risk (n=3,207), RAG-LLM identified 62.7ℒ% (2,010) as high risk, making them newly eligible for anticoagulant therapy. Even after accounting for high bleeding risk, 70.5ℒ% (1,416/2,010) of these would remain candidates for treatment. Among patients with structured evidence of anticoagulation use (n=18,512), RAG-LLM flagged 57.6ℒ% (10,664) as high bleeding risk (HAS-BLEDℒ≥ℒ3), indicating the need for therapy reassessment (**eTable 13**).

## Discussion

In this study, we developed a novel interoperable RAG-LLM system that extracts complex clinical information from structured and unstructured data across non-standardized EHRs. We demonstrated its effectiveness in abstracting risk factors for CHA DS-VASc and HAS-BLED in patients with AF. Compared with traditional rule-based abstraction using structured data alone, the RAG-LLM demonstrated substantially higher accuracy, recall, and efficiency in extracting clinically relevant risk factors, closely matching expert manual annotation while offering over 20-fold improvement in processing time. When deployed across a large real-world cohort of patients with AF, RAG-LLM reclassified 62.1% of individuals initially labeled as low or intermediate stroke risk to high risk, making them eligible for anticoagulation. For bleeding risk, 5.5% of patients categorized as low risk by structured data were reclassified as high risk with RAG-LLM, providing a more accurate assessment of safety of anticoagulation therapy. These findings highlight the potential of RAG-LLM to enhance quality measurement and support scalable, data-driven clinical decision-making.

The RAG-LLM framework addresses critical limitations of previous clinical approaches by enabling scalable abstraction of heterogeneous EHR data elements. Traditional rule-based as well as NLP pipelines are constrained by task-specific development and manual rule creation. They also do not automatically unify structured data with narrative notes, and are difficult to maintain as definitions evolve, which limits their generalizability across tasks.^20^ This precludes their translation across sites and EHR platforms, typically requiring extensive, site-specific customization and re-validation.^20,21^ In contrast, RAG-LLM natively integrates structured and unstructured data without requiring manually codified rules, utilizing a modular retrieval-and-generation architecture that adapts to new phenotypes with minimal changes, thereby supporting cross-system deployment. This approach offers a more efficient and portable solution than traditional NLP approaches.^22–26^

Beyond its technical performance, the RAG-LLM system demonstrated meaningful clinical utility by identifying patients eligible for anticoagulation who would have been missed using methods based on structured data extraction alone. By capturing nuanced clinical details embedded in unstructured documentation, the system enhances the completeness and accuracy of risk assessments.^27–29^ These can also be performed in near real-time as care is delivered, enabling quality improvement interventions to directly follow from observed instances of departure from the standard of care.^16,30^ Moreover, the system’s explainable framework, delivering traceable outputs anchored to source documentation, addresses interpretability and auditability.^31,32^ Another key strength of RAG systems lies in their capacity for dynamic knowledge integration. Rather than embedding information statically within an LLM during training, RAG can be based on any relevant external content, such as clinical practice guidelines or institutional policies, that can be used as a modification of the query.^33,34^

Of note, this study has limitations that merit consideration. First, the study did not include the subset of individuals with incomplete EHR data, specifically those who lacked any clinical notes. This may have introduced selection toward patients with complete documentation and higher healthcare utilization. However, a vast majority of records have at least some clinical documentation, and this would also be an essential component to infer these risk scores for even manual abstraction. Second, the cohort may be enriched for patients already receiving anticoagulation and with more thoroughly documented risk factors, potentially limiting generalizability to newly diagnosed or less-engaged populations. Third, while our expert-annotated reference standard was strengthened by having a clinical group review all discrepancies, manual chart review is subjective, particularly for complex or ambiguously documented clinical features. This subjectivity introduces the possibility of misclassification, which may affect the precision of the reference standard and, in turn, influence the evaluation of model performance. Fourth, the retrospective nature of this analysis may limit our understanding of real-world implementation challenges, but our approach lends itself to pragmatic implementation without requiring further adaptation.

## Conclusion

A multimodal RAG-LLM framework accurately and efficiently abstracts clinical variables from structured and unstructured EHR data. In the context of stroke and bleeding risk stratification, the framework substantially improves assessment of CHA DS-VASc and HAS-BLED scores, supporting more informed recommendations for anticoagulation and advancing the quality of guideline-concordant care. By integrating heterogeneous data sources, RAG-LLM offers a scalable, interoperable solution for improving clinical quality measurement efficiency and precision.

## Data Sharing Statement

The data used for this study from YNHHS cannot be publicly shared, as it represents protected health information and is not permitted under Yale’s IRB approval for the study. The MIMIC-IV cohort has an application for access at https://physionet.org/content/mimiciv/3.0/.

## Funding

Mr. Adejumo is supported by the National Heart Lung and Blood Institute of the National Institutes of Health under award F30HL176149. Dr. Khera is supported by the National Heart Lung and Blood Institute of the National Institutes of Health (under awards R01AG089981, R01HL167858, and K23HL153775) and the Doris Duke Charitable Foundation (under award 2022060). Dr. Thangaraj was supported by the National Institutes of Health under award T32HL155000. Dr. Croon is supported by University of Amsterdam Research Priority Agenda Program AI for Health Decision-Making. The funding organizations had no role in the design and conduct of the study.

## Disclosures

Dr. Khera is an Associate Editor of JAMA. He also receives research support, through Yale, from Bristol-Myers Squibb, Novo Nordisk, and BridgeBio. He is a coinventor of U.S. Pending Patent Applications WO2023230345A1, US20220336048A1, 63/484,426, 63/508,315, 63/580,137, 63/606,203, 63/619,241, 63/562,335, and 63/346,610. He is a co-founder of Ensight-AI, Inc. and Evidence2Health, health platforms to improve cardiovascular diagnosis and evidence-based cardiovascular care. Dr. Thangaraj is a coinventor of a provisional patent (63/606,203). Philip Croon is former owner and still advisor of DGTL Health B.V. The other authors report no relevant disclosures.

## Data Availability

The Yale Institutional Review Board reviewed the study and waived the need for informed consent due to the retrospective nature of the analysis. This study adheres to the TRIPOD (Transparent Reporting of a multivariable prediction model for Individual Prognosis or Diagnosis) reporting guidelines (eTable 1).16

https://physionet.org/content/mimiciv/3.0/

## Notes

### Author Declarations

The Yale Institutional Review Board reviewed the study, approved the protocol, and waived the need for informed consent as this is a secondary analysis of existing data.

### Summary of Updates

This version of the manuscript contains an updated Introduction, Methods, Results, Conclusion, and Supplemental Files. This version contains complete results for CHADS-VASC and HASBLED for YNHHS.

## References

1. Chassin, M. R. & Galvin, R. W. The urgent need to improve health care quality. Institute of Medicine National Roundtable on Health Care Quality. JAMA 280, 1000–1005 (1998).

2. Kumar, A., Wang, H., Muir, K. W., Mishra, V. & Engelhard, M. A cross-sectional study of GPT-4– based plain language translation of clinical notes to improve patient comprehension of disease course and management. NEJM AI 2, (2025).

3. Saraswathula, A. et al. The volume and cost of quality metric reporting. JAMA 329, 1840–1847 (2023).

4. Casalino, L. P. et al. US physician practices spend more than $15.4 billion annually to report quality measures. Health Aff. (Millwood) 35, 401–406 (2016).

5. Wang, Y. et al. Clinical information extraction applications: A literature review. J. Biomed. Inform. 77, 34–49 (2018).

6. Carrell, D. S. et al. Challenges in adapting existing clinical natural language processing systems to multiple, diverse health care settings. J. Am. Med. Inform. Assoc. 24, 986–991 (2017).

7. Singhal, K. et al. Large language models encode clinical knowledge. Nature 620, 172–180 (2023).

8. Boussina, A., et al. Large language models for more efficient reporting of hospital quality measures. NEJM AI 1, (2024).

9. Alkaissi, H. & McFarlane, S. I. Artificial hallucinations in ChatGPT: Implications in scientific writing. Cureus (2023) doi:10.7759/cureus.35179.

10. Gilbert, S., Kather, J. N. & Hogan, A. Augmented non-hallucinating large language models as medical information curators. NPJ Digit. Med. 7, 100 (2024).

11. Liu, S., McCoy, A. B. & Wright, A. Improving large language model applications in biomedicine with retrieval-augmented generation: a systematic review, meta-analysis, and clinical development guidelines. J. Am. Med. Inform. Assoc. 32, 605–615 (2025).

12. Béchard, P. & Ayala, O. M. Reducing hallucination in structured outputs via Retrieval-Augmented Generation. arXiv [cs.LG] (2024).

13. Lip, G. Y. H., Nieuwlaat, R., Pisters, R., Lane, D. A. & Crijns, H. J. G. M. Refining clinical risk stratification for predicting stroke and thromboembolism in atrial fibrillation using a novel risk factor-based approach: the euro heart survey on atrial fibrillation. Chest 137, 263–272 (2010).

14. Olesen, J. B. et al. Validation of risk stratification schemes for predicting stroke and thromboembolism in patients with atrial fibrillation: nationwide cohort study. BMJ 342, d124 (2011).

15. Pisters, R. et al. A novel user-friendly score (HAS-BLED) to assess 1-year risk of major bleeding in patients with atrial fibrillation: the Euro Heart Survey. Chest 138, 1093–1100 (2010).

16. Gallifant, J. et al. The TRIPOD-LLM reporting guideline for studies using large language models. Nat. Med. 31, 60–69 (2025).

17. Johnson, A. E. W. et al. MIMIC-IV, a freely accessible electronic health record dataset. Sci. Data 10, 1 (2023).

18. Lip, G. Y. H. et al. Antithrombotic therapy for atrial fibrillation: CHEST guideline and expert panel report. Chest 154, 1121–1201 (2018).

19. Johnson, L. S. et al. Residual stroke risk among patients with atrial fibrillation prescribed oral anticoagulants: A patient-level meta-analysis from COMBINE AF. J. Am. Heart Assoc. 13, e034758 (2024).

20. Garcia, B. T. et al. Improving automated deep phenotyping through large language models using retrieval-augmented generation. Genome Med. 17, 91 (2025).

21. Amugongo, L. M., Mascheroni, P., Brooks, S., Doering, S. & Seidel, J. Retrieval augmented generation for large language models in healthcare: A systematic review. *PLOS Digit*. Health 4, e0000877 (2025).

22. Li, F., et al. AI-aided dynamic prediction of bleeding and ischemic risk after coronary stenting and subsequent DAPT. bioRxiv (2022) doi:10.1101/2022.02.05.22270508.

23. Elkin, P. L. et al. Using artificial intelligence with natural language processing to combine electronic health record’s structured and free text data to identify nonvalvular atrial fibrillation to decrease strokes and death: Evaluation and case-control study. J. Med. Internet Res. 23, e28946 (2021).

24. Shung, D., Simonov, M., Gentry, M., Au, B. & Laine, L. Machine learning to predict outcomes in patients with acute gastrointestinal bleeding: A systematic review. Dig. Dis. Sci. 64, 2078–2087 (2019).

25. Taggart, M. et al. Abstract 11: Development and comparison of two natural language processing methods for identifying bleeding events in clinical text. Circ. Cardiovasc. Qual. Outcomes 11, (2018).

26. Liang, J. J. et al. Towards generalizable methods for automating risk score calculation. in *Proceedings of the 21st Workshop on Biomedical Language Processing* (Association for Computational Linguistics, Stroudsburg, PA, USA, 2022). doi:10.18653/v1/2022.bionlp-1.42.

27. Zhang, Y. et al. High-throughput phenotyping with electronic medical record data using a common semi-supervised approach (PheCAP). Nat. Protoc. 14, 3426–3444 (2019).

28. Nargesi, A. A. et al. Automated identification of heart failure with reduced ejection fraction using deep learning-based natural language processing. JACC Heart Fail. 13, 75–87 (2024).

29. Cunningham, J. W. et al. Natural language processing for adjudication of heart failure in the electronic health record. JACC Heart Fail. 11, 852–854 (2023).

30. Amann, J. et al. To explain or not to explain?-Artificial intelligence explainability in clinical decision support systems. PLOS Digit. Health 1, e0000016 (2022).

31. Mohapatra, R. K., Jolly, L. & Dakua, S. P. Advancing explainable AI in healthcare: Necessity, progress, and future directions. Comput. Biol. Chem. 108599 (2025).

32. Alkhanbouli, R., Matar Abdulla Almadhaani, H., Alhosani, F. & Simsekler, M. C. E. The role of explainable artificial intelligence in disease prediction: a systematic literature review and future research directions. BMC Med. Inform. Decis. Mak. 25, 110 (2025).

33. Yang, R. et al. Retrieval-augmented generation for generative artificial intelligence in health care. Npj Health Syst. 2, 1–5 (2025).

34. Thirunavukarasu, A. J. et al. Large language models in medicine. Nat. Med. 29, 1930–1940 (2023).

35. Unlu, O. et al. Retrieval Augmented Generation enabled generative pre-trained transformer 4 (GPT-4) performance for clinical trial screening. medRxiv (2024) doi:10.1101/2024.02.08.24302376.

36. Zhang, G. et al. Leveraging long context in retrieval augmented language models for medical question answering. NPJ Digit. Med. 8, 239 (2025).

