## Supplemental File for "Evaluation of Care Quality for Atrial Fibrillation Across Non-Interoperable Electronic Health Record Data using a Retrieval-Augmented Generation-enabled Large Language Model"

**eMethods**

**Software and Libraries**

A Python-based computational environment was used for all query design, annotation, and retrieval tasks. The work primarily utilized Pandas for data ingestion, cleaning, and preprocessing of both structured and unstructured patient data, as well as NumPy for numerical operations such as array manipulation and basic statistical analysis. All embedded text segments were stored and retrieved using Chroma as the vector database, allowing efficient similarity-based lookups. Domain-agnostic yet contextually rich embeddings of clinical text were generated using NV-embed. After an initial cosine similarity stage, Cross-Encoder re-ranking models were applied for more precise retrieval by comparing the full context of queries and candidate text segments. Finally, Qwen‑3‑32B served as the LLM for synthesizing final outputs.

**Data Pre-processing and Schema Mapping**

Data pre-processing took place at both the structured and unstructured levels. First, we extracted patient demographics, diagnosis codes, procedure codes, laboratory data, and selected clinical measurements from the EHR's structured fields. These data were stored in relational tables that mapped each patient to their recorded encounters, facilitating cross-referencing of codes with text-derived evidence.

**Automated Schema Mapping**

The RAG-LLM framework implements a novel two-stage approach for automated schema mapping, eliminating the need for manual curation. This system processes heterogeneous healthcare data through intelligent table classification followed by context-aware column mapping. We engineered an LLM-based classifier that analyzes data content patterns to categorize each table into one of nine standard healthcare data types: patient demographics, diagnoses, medications, laboratory results, procedures, encounters, vital signs, clinical notes, or reference dictionaries. The classifier examines column names, data patterns, uniqueness ratios, and sample values to determine table type with high confidence. Once classified, tables undergo automated column mapping specific to their type. The system uses an LLM to map source columns to standardized fields based on semantic understanding rather than rigid rules.

**Vectorization Process**

After mapping, both structured tables and unstructured text undergo vectorization. Structured data is serialized with semantic field names, while clinical notes are segmented at the sentence level using the Natural Language Toolkit. For the unstructured data, clinical notes such as provider narratives, discharge summaries, and progress notes were parsed to retain sections most relevant to cardiovascular risk factors. We leveraged a clinical text sentencizer from MedSpacy to split clinical text into individual sentence chunks. This sentence-level tokenization increased the precision of retrieval because it restricted the system to returning only the most pertinent text snippets, rather than entire paragraphs or sections. By pinpointing specific sentences where risk factors were documented, we minimized the retrieval of extraneous or irrelevant textual content, improving overall efficiency in the subsequent retrieval steps. This creates a unified vector space where semantically related information clusters together regardless of original format, enabling the RAG system to retrieve relevant evidence across data modalities seamlessly.

**Query Development and Clinical Validation Methodology**

Our approach involved designing hybrid structured–unstructured queries to identify risk factors for CHA₂DS₂-VASc and HAS-BLED. Each risk factor was linked to a standardized query that referenced both codes and narrative descriptors. On the structured side, we included ICD-9 and ICD-10 codes, and lab values indicative of risk. On the unstructured side, we incorporated natural-language definitions, synonyms, and clinical descriptors that commonly appear in clinical notes.

The query construction process integrated clinical expertise to ensure alignment with contemporary cardiovascular practice patterns (**Supplementary Figure 2**). Initially, an expert panel consisting of two clinicians and one cardiologist reviewed the original CHA₂DS₂-VASc and HAS-BLED validation studies alongside current ACC/AHA guideline recommendations. This panel systematically mapped each risk factor to both its canonical presentations and the heterogeneous documentation patterns observed in clinical practice. For structured data mapping, clinicians reviewed ICD-9/10 code sets for comprehensiveness, supplementing with clinically equivalent diagnoses that might be coded differently across institutions. For unstructured data, clinicians annotated 10 representative clinical notes to identify discipline-specific terminology, common abbreviations, contextual modifiers, and temporal qualifiers that substantially alter clinical interpretation.

The panel adjudicated discrepancies through a modified Delphi approach. Following initial query construction, we employed a clinician-supervised validation process on a development cohort, where retrieval results were manually verified against full chart review. This refinement phase calibrated similarity thresholds and optimized the cross-encoder reranking function to prioritize clinically significant evidence patterns.

**Error Analysis Methodology**

Following completion of independent annotations and consensus adjudication, we systematically analyzed all discordant cases between expert reviewers and RAG-LLM determinations. Each discordance was categorized by the consensus panel into predefined error types:

**Expert Annotation Error Categories:**

1. Laboratory value oversight - Missing values marginally exceeding thresholds
2. Subspecialty documentation gaps - Information in specialized consultation notes
3. Complex pathology interpretation - Nuanced clinical conditions requiring expertise

**RAG-LLM Error Categories:**

1. Contextual misinterpretation - Isolated findings interpreted without full context
2. Temporal disambiguation - Difficulty distinguishing timing of events
3. Acute vs chronic distinction - Challenges differentiating condition duration

The consensus panel reviewed each discordant case, identified the error source (expert vs RAG-LLM), and assigned it to the most appropriate category. Percentages were calculated based on total errors within each cohort.

**Model Architecture and Technical Specifications**

The information processing pipeline began with converting heterogeneous clinical documentation into a standardized vector space using NV-embed, a general-purpose neural embedding model that transformed diverse documentation styles into dense representations.^21^ The system then indexed these vectors in a specialized database optimized for similarity retrieval using cosine similarity measurements. For each clinical query, the system selected the top 20 most relevant text segments based on semantic similarity scores, which were combined with the standardized query and passed to our analytical engine. This analytical component utilized the 32-billion parameter Qwen 3 model as the reasoning engine, selected for its decoder architecture that enables explicit reasoning pathways with transparent documentation of clinical decision logic.^22–24^ The final structured assessment, along with supporting evidence and reasoning pathway, was stored in a queryable database that enabled full transparency into both the label determination and its corresponding evidence. For each query, the model evaluates retrieved information against standardized clinical definitions, generating structured outputs with supporting evidence that maintains provenance to the original documentation.

**Integration Into the RAG Pipeline**

Once the annotated text segments were finalized, they were used to enrich patient-level data by integrating structured elements (coded diagnoses and clinical measurements) with unstructured evidence drawn from the notes. We assembled all relevant textual and coded indications for each risk factor, consolidating them into a single representation for every patient.

The final extraction and scoring took place through the Llama 3.3 LLM, which generated a binary or multiclass assessment of each risk factor (present, absent, or degree of severity) and appended a brief text excerpt justifying its classification. With these assessments, we computed each patient’s CHA₂DS₂-VASc and HAS-BLED scores by combining all risk factor statuses. In addition to the final classification, the RAG workflow retained chain-of-thought reasoning for internal auditing, though these intermediate reasoning steps were not shared externally. This multi-stage pipeline significantly improved our ability to locate and interpret risk factors that might otherwise remain scattered or obscured in the complexities of clinical documentation.

**Structured Data Extraction Methodology**

For comparison with RAG-LLM performance, we implemented rule-based abstraction from structured data sources. This approach used:

- Direct ICD-9 and ICD-10 code matching using the comprehensive code lists provided in Supplementary Table 4
- Laboratory value extraction with threshold-based classification (e.g., creatinine >2.26 mg/dL for renal disease)
- Medication identification through exact name matching and therapeutic class groupings
- Boolean logic determining risk factor presence based on any qualifying criterion

The structured approach could not access narrative text, clinical impressions, or contextual modifiers that might alter interpretation. This limitation was particularly evident for conditions requiring clinical judgment or temporal context.

**Manual Annotation Guidelines for CHA₂DS₂-VASc and HAS-BLED Risk Factors**

**Overview**

The primary objective of our manual annotation process was to establish a gold standard for the identification of stroke risk (CHA₂DS₂-VASc) and bleeding risk (HAS-BLED) factors in patients with atrial fibrillation. Annotators were instructed to systematically review both structured data elements (diagnostic codes, laboratory values, medication lists) and unstructured clinical documentation to determine the presence or absence of each risk factor according to standardized clinical criteria.

**Annotation Timeline and Workflow**

The annotation process followed a standardized workflow:

1. Initial review of structured data elements (5-7 minutes)
2. Comprehensive review of clinical notes (10-15 minutes)
3. Final determination and documentation (3-5 minutes)
4. Total average time: 12 minutes per patient (range: 10-15 minutes)

Annotators (PMT, PMC) recorded time per batch of 10 patients using an automated timing system. Inter-rater reliability was assessed on the first 50 patients from each site, achieving substantial agreement (κ=0.82). For the 15% of cases with disagreements, the consensus panel spent an additional 10-15 minutes per case in structured adjudication.

**General Annotation Principles**

For each risk factor, annotators were instructed to:

1. Consider both structured and unstructured data sources
2. Apply clinical judgment when evidence was conflicting
3. Prioritize current clinical status over historical mentions unless specifically relevant
4. Consider temporal context for all findings
5. Follow the principle that chronic conditions remain present even if well-controlled

**1. Congestive Heart Failure**

**Qualifying Criteria:**

- Direct documentation of "CHF," "heart failure," "HFpEF," "HFrEF," "systolic heart failure," or "diastolic heart failure"
- ICD-10 codes from the I50.x series
- Echocardiographic evidence of left ventricular dysfunction (LVEF ≤ 45%)
- Clinical documentation of volume overload attributed to cardiac dysfunction
- Active management with heart failure medications specifically for heart failure

**Non-Qualifying Criteria:**

- Heart failure explicitly described as resolved with normal current cardiac function
- Documentation of "rule out CHF" or "CHF unlikely" without subsequent confirmation
- Isolated mention of edema or dyspnea attributed to non-cardiac causes
- Heart failure mentioned only in family history or risk assessment
- Normal ejection fraction without other signs of heart failure

**Notes and Edge Cases:**

- For patients with borderline ejection fraction (40-45%), annotators were instructed to look for supporting clinical evidence such as diuretic use or documented volume overload
- For patients with improved ejection fraction following treatment, heart failure was still considered present if the patient remained on heart failure medications
- When conflicting EF values were present, annotators were instructed to use the most recent values unless acute illness explained the discrepancy

**2. Hypertension**

**Qualifying Criteria:**

- Direct diagnosis of "hypertension" or "HTN" in problem list or clinical notes
- ICD-10 codes from the I10-I16 series
- Antihypertensive medication prescription with explicit indication for hypertension
- Documentation of target organ damage specifically attributed to hypertension

**Non-Qualifying Criteria:**

- Isolated elevated blood pressure readings without diagnosis of hypertension
- White coat hypertension explicitly noted as the only form of hypertension
- Hypertension documented as completely resolved following an intervention (e.g., renal artery stenosis repair)
- Transient hypertension during acute illness without continuing diagnosis

**Notes and Edge Cases:**

- Controlled hypertension still qualified as hypertension for scoring purposes
- For notations of "history of hypertension" without clear indication of resolution, hypertension was considered present
- For patients on antihypertensives with dual indications (e.g., beta-blockers for both HTN and rate control), annotators were instructed to look for explicit HTN diagnosis

**3. Diabetes**

**Qualifying Criteria:**

- Documented diagnosis of Type 1 or Type 2 diabetes mellitus
- ICD-10 codes from the E08-E13 series
- Current prescription of insulin or oral hypoglycemic agents
- Laboratory values meeting diagnostic criteria:
  - HbA1c ≥ 6.5% (48 mmol/mol)
  - Fasting Plasma Glucose ≥ 126 mg/dL (7.0 mmol/L)
  - Random Plasma Glucose ≥ 200 mg/dL (11.1 mmol/L) with symptoms

**Non-Qualifying Criteria:**

- Prediabetes or impaired glucose tolerance without diabetes diagnosis
- Gestational diabetes without subsequent Type 2 diabetes diagnosis
- "Rule out diabetes" without confirmed diagnosis
- Isolated elevated glucose readings during acute illness or steroid treatment without diabetes diagnosis

**Notes and Edge Cases:**

- Diet-controlled diabetes was considered present even without medication
- For patients with "borderline diabetes," annotators were instructed to look for any explicit diabetes diagnosis elsewhere in the record
- When blood glucose values were inconsistently elevated, annotators were to prioritize HbA1c as the more reliable indicator

**4. Stroke/TIA/Thromboembolism**

**Qualifying Criteria:**

- Documented history of ischemic stroke, hemorrhagic stroke, or TIA
- ICD-10 codes: I63.x (ischemic stroke), I61.x (hemorrhagic stroke), G45.x (TIA)
- Radiographic evidence of previous stroke or cerebral infarction
- Documented systemic thromboembolism (e.g., pulmonary embolism, arterial embolism)
- Clear documentation of neurological deficits attributed to cerebrovascular event

**Non-Qualifying Criteria:**

- Stroke or TIA specifically "ruled out" after evaluation
- Neurological symptoms attributed to other causes (migraine, seizure)
- Documentation of "stroke-like symptoms" without confirmed diagnosis
- Family history of stroke without personal history

**Notes and Edge Cases:**

- For unclear documentation (e.g., "possible TIA"), annotators were instructed to look for supporting evidence like neurology consultation or antiplatelet initiation
- Remote events still qualified, regardless of time since occurrence
- When determining between TIA and stroke, annotators were to use the final diagnosis rather than initial presentation

**5. Vascular Disease**

**Qualifying Criteria:**

- Documented coronary artery disease, previous myocardial infarction, or coronary intervention
- ICD-10 codes: I21.x (acute MI), I25.2 (old MI), I25.1x (coronary atherosclerosis)
- Peripheral arterial disease with symptoms or requiring intervention (ICD-10: I70.2x, I73.8)
- Aortic plaque or aortic aneurysm
- Previous vascular interventions:
  - Coronary artery bypass grafting (CABG)
  - Percutaneous coronary intervention (PCI)
  - Peripheral vascular bypass or angioplasty

**Non-Qualifying Criteria:**

- Non-obstructive coronary artery disease explicitly described as minimal or non-significant
- Venous disease (e.g., DVT, venous insufficiency) without arterial involvement
- Family history of vascular disease without personal history
- Risk factors for vascular disease without established diagnosis

**Notes and Edge Cases:**

- For patients with coronary calcium scores, a score >400 was considered evidence of vascular disease
- For borderline cases with terms like "mild coronary disease," annotators were instructed to look for corresponding medical therapy (e.g., statins, antiplatelets)
- Vascular disease remained present even if successfully treated with interventions

**6. Abnormal Renal Function**

**Qualifying Criteria:**

- Documented chronic kidney disease (CKD), end-stage renal disease (ESRD), or renal failure
- ICD-10 codes from the N18.x series
- Current or prior dialysis or kidney transplant
- Laboratory values:
  - Single creatinine > 2.26 mg/dL (200 µmol/L)
  - Sustained creatinine > 1.5 mg/dL on multiple readings
  - eGFR < 30 mL/min consistently documented

**Non-Qualifying Criteria:**

- Acute kidney injury (AKI) that fully resolved
- Mild renal impairment described as clinically insignificant
- Isolated abnormal lab value without clinical correlation
- Renal cysts or anatomical abnormalities without functional impairment

**Notes and Edge Cases:**

- For patients with fluctuating renal function, chronic pattern took precedence over isolated readings
- For patients receiving nephrology care, annotators were instructed to consider this as supporting evidence even if lab values were borderline
- Medication adjustments specifically for renal function were considered supporting evidence

**7. Abnormal Liver Function**

**Qualifying Criteria:**

- Documented cirrhosis (any etiology), chronic liver disease, or hepatic dysfunction
- ICD-10 codes from the K70-K77 series
- Laboratory values indicating severe liver dysfunction:
  - Total bilirubin > 2x upper limit of normal (typically >2.4 mg/dL) WITH
  - At least one of:
    - AST > 3x upper limit of normal (>120 IU/L)
    - ALT > 3x upper limit of normal (>120 IU/L for men, >90 IU/L for women)
    - Alkaline phosphatase > 3x upper limit of normal (>120 IU/L)

**Non-Qualifying Criteria:**

- Isolated transaminitis that resolved
- Fatty liver without evidence of dysfunction
- Mild hepatic steatosis without laboratory abnormalities
- Hepatitis with full recovery and normal current function

**Notes and Edge Cases:**

- For patients with a history of alcohol abuse, annotators were instructed to look carefully for evidence of liver dysfunction even if not explicitly documented
- Portal hypertension, ascites, or varices were considered strong evidence of liver dysfunction
- For chronic viral hepatitis, evidence of fibrosis or cirrhosis was required to qualify

**8. History of Bleeding**

**Qualifying Criteria:**

- Documented history of:
  - Intracranial hemorrhage (ICD-10: I60-I62)
  - Gastrointestinal bleeding (ICD-10: K92.2)
  - Major bleeding requiring hospitalization or transfusion
  - Bleeding causing significant hemoglobin drop (>2 g/dL)
  - Other significant bleeding (retroperitoneal, intra-articular, pericardial)
- Diagnosed bleeding disorders: coagulopathy, thrombocytopenia, von Willebrand disease
- ICD-10 code D68.x diagnoses

**Non-Qualifying Criteria:**

- Minor bleeding (e.g., epistaxis, gum bleeding) that resolved without intervention
- Bleeding events clearly related to trauma without underlying predisposition
- Family history of bleeding disorders without personal manifestation
- Mention of bleeding risk without documented bleeding events

**Notes and Edge Cases:**

- For patients with recurrent minor bleeding, annotators were instructed to consider the pattern and frequency
- Remote bleeding events (>5 years ago) still qualified if significant
- Post-procedural bleeding was included only if excessive or requiring intervention beyond standard care
- For anemia due to chronic blood loss, source documentation was required to qualify as bleeding history

**9. Labile INR**

**Qualifying Criteria:**

- Explicit documentation of "labile INR," "unstable INR," or "poor INR control"
- For patients on warfarin:
  - Documented time in therapeutic range (TTR) < 60%
  - ≥2 INR values outside therapeutic range (2.0-3.0) within 6 months
  - ≥2 INR values > 5.0 within 6 months
  - Frequent warfarin dose adjustments (≥2 in a month)
- For patients not on warfarin:
  - ≥2 INR values > 1.5 without anticoagulation therapy
  - Unexplained elevated INR in absence of anticoagulants

**Non-Qualifying Criteria:**

- Isolated out-of-range INR measurements with documented explanation (e.g., medication interaction)
- INR fluctuations during warfarin initiation phase
- Intentional supra-therapeutic anticoagulation for specific indication
- Abnormal INR due to liver disease (counted under liver dysfunction only)

**Notes and Edge Cases:**

- For patients recently started on warfarin, annotators were instructed to disregard initial adjustment period
- For patients with documented non-compliance, annotators were instructed to consider this as evidence of labile INR
- Medical record documentation quality significantly affected ability to assess this criterion

**10. Bleeding Medications**

**Qualifying Criteria:**

- Current use of:
  - Antiplatelet medications: Aspirin (any dose), clopidogrel, prasugrel, ticagrelor
  - Dual antiplatelet therapy (DAPT)
  - NSAIDs: Regular use of ibuprofen, naproxen, diclofenac, etc.
  - Other medications with significant bleeding risk

**Non-Qualifying Criteria:**

- Previous use of bleeding medications that were discontinued
- As-needed (PRN) use of NSAIDs without regular schedule
- Low-dose aspirin (81mg) used for cardiovascular protection alone
- Anticoagulants (these were considered separately for clinical decision-making)

**Notes and Edge Cases:**

- For patients on multiple bleeding risk medications, each agent was documented separately
- For patients on anticoagulants plus antiplatelets, this was considered a particularly high-risk combination
- Temporary interruptions in therapy were disregarded if the medication was being resumed

**11. Alcohol Use**

**Qualifying Criteria:**

- Documented consumption ≥8 drinks per week
- Documentation of "heavy," "excessive," or "significant" alcohol use
- Screening results:
  - AUDIT-C score ≥4 for men or ≥3 for women
  - Other alcohol screening tool results indicating significant use
- Documented alcohol use disorder or alcoholism

**Non-Qualifying Criteria:**

- Social or occasional drinking explicitly described as modest
- Remote history of alcohol abuse with documented sobriety
- Documentation of "non-drinker" or "rare/occasional alcohol use"
- Alcohol use without quantification when described as minimal

**Notes and Edge Cases:**

- When alcohol use was documented without specific quantities, annotators were instructed to look for descriptive terms suggesting excess
- For inconsistent documentation, annotators were instructed to use the most recent assessment
- For patients with alcohol-related medical complications, significant use was presumed even without specific quantification

**12. Uncontrolled Hypertension**

**Qualifying Criteria:**

- Explicit documentation of "uncontrolled hypertension," "poorly controlled hypertension," or "resistant hypertension"
- Blood pressure measurements:
  - Systolic BP > 160 mmHg on ≥2 separate occasions within past 6 months
  - Consistently elevated BP despite being on ≥3 antihypertensive medications
  - Documentation of failure to reach BP target despite medication adherence

**Non-Qualifying Criteria:**

- Isolated elevated BP readings during acute illness, pain, or stress
- White coat hypertension with normal home measurements
- Well-controlled hypertension with occasional elevations
- Elevated readings in non-hypertensive patients

**Notes and Edge Cases:**

- Annotators were instructed to consider the overall pattern of BP control rather than isolated readings
- For patients with diabetes or CKD, stricter BP targets were considered (>140/90 mmHg could indicate poor control)
- For patients with documented medication non-adherence, hypertension control was assessed based on periods of proper medication use when available

**eTable 1. Transparent Reporting of a Multivariable Prediction Model for Individual Prognosis or Diagnosis (TRIPOD) Checklist for Reporting Guideline Compliance.**

| Section/Topic | Item | Checklist Item | Page | Reported |
| --- | --- | --- | --- | --- |
| Title and Abstract |  |  |  |  |
| Title | 1 | Identify the study as developing and/or validating a multivariable prediction model, the target population, and the outcome to be predicted. | 1 | Yes |
| Abstract | 2 | Provide a summary of objectives, study design, setting, participants, sample size, predictors, outcome, statistical analysis, results, and conclusions. | 1 | Yes |
| Introduction |  |  |  |  |
| Background and objectives | 3a | Explain the medical context (including whether diagnostic or prognostic) and rationale for developing or validating the multivariable prediction model, including references to existing models. | 2-3 | Yes |
|  | 3b | Specify the objectives, including whether the study describes the development or validation of the model or both. | 3 | Yes |
| Methods |  |  |  |  |
| Source of data | 4a | Describe the study design or source of data (e.g., randomized trial, cohort, or registry data), separately for the development and validation datasets, if applicable. | 4-5 | Yes |
|  | 4b | Specify the key study dates, including start of accrual; end of accrual; and, if applicable, end of follow-up. | 4-5 | Yes |
| Participants | 5a | Specify key elements of the study setting (e.g., primary care, secondary care, general population) including number and location of centres. | 4-5 | Yes |
|  | 5b | Describe eligibility criteria for participants. | 5 | Yes |
|  | 5c | Give details of treatments received, if relevant. | N/A | N/A |
| Outcome | 6a | Clearly define the outcome that is predicted by the prediction model, including how and when assessed. | 5-6 | Yes |
|  | 6b | Report any actions to blind assessment of the outcome to be predicted. | 7 | Yes |
| Predictors | 7a | Clearly define all predictors used in developing or validating the multivariable prediction model, including how and when they were measured. | 6-7 | Yes |
|  | 7b | Report any actions to blind assessment of predictors for the outcome and other predictors. | 7 | Yes |
| Sample size | 8 | Explain how the study size was arrived at. | 5 | Yes |
| Missing data | 9 | Describe how missing data were handled (e.g., complete-case analysis, single imputation, multiple imputation) with details of any imputation method. | 5 | Partial |
| Statistical analysis methods | 10a | Describe how predictors were handled in the analyses. | 7-8 | Yes |
|  | 10b | Specify type of model, all model-building procedures (including any predictor selection), and method for internal validation. | 6-8 | Yes |
|  | 10c | For validation, describe how the predictions were calculated. | 8-9 | Yes |
|  | 10d | Specify all measures used to assess model performance and, if relevant, to compare multiple models. | 9-10 | Yes |
|  | 10e | Describe any model updating (e.g., recalibration) arising from the validation, if done. | N/A | N/A |
| Risk groups | 11 | Provide details on how risk groups were created, if done. | 9 | Yes |
| Development vs. validation | 12 | For validation, identify any differences from the development data in setting, eligibility criteria, outcome, and predictors. | 5 | Yes |
| Results |  |  |  |  |
| Participants | 13a | Describe the flow of participants through the study, including the number of participants with and without the outcome and, if applicable, a summary of the follow-up time. A diagram may be helpful. | 10-11 | Yes |
|  | 13b | Describe the characteristics of the participants (basic demographics, clinical features, available predictors), including the number of participants with missing data for predictors and outcome. | 10-11 | Yes |
|  | 13c | For validation, show a comparison with the development data of the distribution of important variables (demographics, predictors and outcome). | 10-11 | Yes |
| Model development | 14a | Specify the number of participants and outcome events in each analysis. | 11-13 | Yes |
|  | 14b | If done, report the unadjusted association between each candidate predictor and outcome. | 12-13 | Partial |
| Model specification | 15a | Present the full prediction model to allow predictions for individuals (i.e., all regression coefficients, and model intercept or baseline survival at a given time point). | N/A | N/A |
|  | 15b | Explain how to the use the prediction model. | N/A | N/A |
| Model performance | 16 | Report performance measures (with CIs) for the prediction model. | 11-12 | Yes |
| Model-updating | 17 | If done, report the results from any model updating (i.e., model specification, model performance). | N/A | N/A |
| Discussion |  |  |  |  |
| Limitations | 18 | Discuss any limitations of the study (such as nonrepresentative sample, few events per predictor, missing data). | 15-16 | Yes |
| Interpretation | 19a | For validation, discuss the results with reference to performance in the development data, and any other validation data. | 14-15 | Yes |
|  | 19b | Give an overall interpretation of the results, considering objectives, limitations, results from similar studies, and other relevant evidence. | 14-16 | Yes |
| Implications | 20 | Discuss the potential clinical use of the model and implications for future research. | 16 | Yes |
| Other Information |  |  |  |  |
| Supplementary information | 21 | Provide information about the availability of supplementary resources, such as study protocol, Web calculator, and datasets. | 17 | Partial |
| Funding | 22 | Give the source of funding and the role of the funders for the present study. | 17 | Yes |

**eTable 2. Standardized Query Criteria for Clinical Risk Factor Identification in Electronic Health Record (EHR) Data.** Detailed specifications of key criteria and instructions used for extracting risk factors from structured and unstructured data.

| **Condition** | **Query Summary (Key Criteria & Instructions)** |
| --- | --- |
| **Hypertension** | **Key Criteria:** Direct mentions of hypertension or “HTN,” ICD-10 I10-I16, documented HTN medications (ACE inhibitors, ARBs, beta blockers, etc.), and supporting evidence (target organ damage, BP monitoring). **Controlled vs. Uncontrolled:** Uncontrolled if explicitly documented (“uncontrolled HTN”), multiple systolic readings >160, medication adjustments for BP control, or nonadherence. **Output:** [Yes/No], [Controlled/Uncontrolled], Evidence quotes. |
| **Congestive Heart Failure** | **Key Criteria:** Mentions of “CHF” or “heart failure,” ICD-10 I50.x, mention of HFpEF/HFrEF. Echo/imaging showing LVEF ≤ 40% or moderate/severe dysfunction. Combination of EF (45–50%) + management indicators (diuretics, volume overload) can also confirm. **Output:** [Yes/No], quoted evidence, explanation of decision logic. |
| **Diabetes Mellitus** | **Key Criteria:** Mentions of “diabetes,” ICD-10 E08-E13, diabetes medications (insulin, oral hypoglycemics), or lab values meeting diagnostic thresholds (HbA1c ≥ 6.5%, FPG ≥ 126 mg/dL). **Output:** [Yes/No], quotes for labs, medications, or notes, plus explanation of whether T1DM or T2DM if specified. |
| **Stroke/TIA/Thromboembolism** | **Key Criteria:** Mentions of “stroke,” “CVA,” “TIA,” ICD-10 codes (e.g., I60.x, I63.x, G45.x). Radiological evidence (CT/MRI) confirming infarction/hemorrhage also qualifies. Thromboembolic events included if specifically documented (ICD-10 I26.x, I74.x). **Output:** [Yes/No], quotes from text, event type (Stroke/TIA/Thromboembolism), and dates if available. |
| **Vascular Disease** | **Key Criteria:** MI/coronary disease (ICD-10 I21.x, I25.x), peripheral arterial disease (I70.2, I73.8), aortic disease (aortic plaque, atherosclerosis, etc.), or any related interventions (CABG, PCI, vascular bypass). **Output:** [Yes/No], disease categories (e.g., MI/Coronary, PAD, Aortic), anatomic locations, and quoted evidence. |
| **Renal Disease** | **Key Criteria:** Chronic kidney disease (CKD), ESRD, ICD-10 N18.x, sustained creatinine elevation (>2.26 mg/dL or >1.5 mg/dL on multiple readings), dialysis or transplant history. **Output:** [Yes/No], key lab values/dates, quotes, plus any nephrology follow-up or medication adjustments. |
| **Liver Disease** | **Key Criteria:** Cirrhosis, ESLD, or chronic liver disease documentation, ICD-10 codes (K70.3, K74.x, etc.), or evidence of related complications (portal HTN, encephalopathy). Radiologic evidence (fibrosis, cirrhosis) also qualifies. **Output:** [Yes/No], event type (cirrhosis, chronic liver disease, other), quotes, and relevant dates. |
| **History of Major Bleeding** | **Key Criteria:** Any documented GI bleed, intracranial hemorrhage, or other severe bleeding; ICD-10 codes referencing bleeding disorders or events. Endoscopy/imaging showing active or past bleeding. **Output:** [Yes/No], event type (GI bleed, intracranial bleed, etc.), quotes, and relevant dates. |
| **Labile INR** | **Key Criteria:** Explicit mention of “labile,” “unstable,” or “poor” INR control, repeated high INR (>5.0) or very low INR (<1.5) within a short timeframe, TTR <60%, frequent dose adjustments. **Output:** [Yes/No], listing of INR values/dates, evidence of instability, and any management changes. |
| **Bleeding Medications** | **Key Criteria:** Documentation of antiplatelet use (Aspirin, P2Y12 inhibitors), NSAIDs, or combination therapy that predisposes to bleeding. **Output:** [Yes/No], list of medications (with dates), quotes, and explanation. |
| **Alcohol Use** | **Key Criteria:** Documented consumption ≥8 drinks/week, screening tools (AUDIT, CAGE) showing heavy use, or explicit mention of “excessive” or “heavy” drinking patterns. **Output:** [Yes/No], direct quotes indicating quantity/timing, and relevant details (e.g., mention of dependence, counseling). |
| **Uncontrolled HTN** | **Key Criteria:** Specific mention of “uncontrolled” or “poorly controlled” hypertension, persistent systolic readings >150 or >140 (depending on comorbidities), recent escalation of BP meds, or non-adherence. **Output:** [Yes/No], quotes, BP readings/dates, explanation of logic, labeled as [Uncontrolled]. |

**eTable 3: Required Data Components and Elements for Multimodal Retrieval-Augmented Generation Framework Implementation.** Patient identification (ID), International Classification of Diseases 9th and 10th Revision (ICD-9/10).

| Data Component | Required Elements |
| --- | --- |
| Patient Demographics | Patient ID, Age, Sex |
| Diagnosis Codes | ICD-9/10 codes with dates |
| Laboratory Values | Test name, result, units, reference range, date |
| Medications | Name, dose, frequency, start/end dates |
| Clinical Notes | Full text, note type, date |
| Query Templates | Risk factor definitions following format in Supplementary Table 3 |

**eTable 4. International Classification of Diseases, 10th Revision (ICD-10) Diagnosis Codes Used for Automated Identification of CHA₂DS₂-VASc and HAS-BLED Risk Factor Components.**

**CHA₂DS₂-VASc Components**

**Congestive Heart Failure**

- **I50.*** (all heart failure codes)
  - I50.1 (left ventricular failure)
  - I50.2* (systolic heart failure)
  - I50.3* (diastolic heart failure)
  - I50.4* (combined systolic and diastolic heart failure)
  - I50.8* (other heart failure)
  - I50.9 (heart failure, unspecified)
- **I11.0** (hypertensive heart disease with heart failure)
- **I13.0, I13.2** (hypertensive heart and chronic kidney disease with heart failure)

**Hypertension**

- **I10** (essential primary hypertension)
- **I11.*** (hypertensive heart disease)
- **I12.*** (hypertensive chronic kidney disease)
- **I13.*** (hypertensive heart and chronic kidney disease)
- **I15.*** (secondary hypertension)
- **I16.*** (hypertensive crisis)
- **I97.3** (postprocedural hypertension)
- **N26.2** (page kidney)

**Diabetes Mellitus**

- *E08.5, E08.6*** (drug or chemical induced diabetes with complications)
- **E09.5*** (drug or chemical induced diabetes with circulatory complications)
- **E10.*** (type 1 diabetes mellitus - all subcodes)
- **E11.*** (type 2 diabetes mellitus - all subcodes)
- **E13.*** (other specified diabetes mellitus - all subcodes)

**Stroke/TIA/Thromboembolism**

- **G45.*** (transient cerebral ischemic attacks and related syndromes)
- **G46.*** (vascular syndromes of brain in cerebrovascular diseases)
- **I26.*** (pulmonary embolism)
- **I60.*** (nontraumatic subarachnoid hemorrhage)
- **I61.*** (nontraumatic intracerebral hemorrhage)
- **I63.*** (cerebral infarction)
- *I67.84, I67.89** (other cerebrovascular disorders)
- **I74.*** (arterial embolism and thrombosis)
- **T80.0XXA** (air embolism following infusion, transfusion and therapeutic injection)
- **T81.718A, T81.72XA** (complication of artery following a procedure)
- **T82.817A, T82.818A** (embolism/thrombosis due to cardiac devices)

**Vascular Disease**

- **I21.*** (acute myocardial infarction)
- **I22.*** (subsequent ST elevation and non-ST elevation myocardial infarction)
- **I25.*** (chronic ischemic heart disease)
  - I25.1* (atherosclerotic heart disease of native coronary artery)
  - I25.2 (old myocardial infarction)
- **I42.*** (cardiomyopathy)
- **I43** (cardiomyopathy in diseases classified elsewhere)
- **I70.*** (atherosclerosis)
  - I70.0 (atherosclerosis of aorta)
  - I70.1 (atherosclerosis of renal artery)
  - I70.2* (atherosclerosis of native arteries of extremities)
  - I70.3*-I70.7* (atherosclerosis of other arteries)
  - I70.8-I70.9* (other and generalized atherosclerosis)
- **I73.1, I73.9** (other peripheral vascular diseases)
- **I79.1, I79.8** (disorders of arteries in diseases classified elsewhere)

**HAS-BLED Components**

**Abnormal Renal Function**

- **N18.*** (chronic kidney disease)
- **N19** (unspecified kidney failure)
- **N26.1, N26.2, N26.9** (unspecified contracted kidney)

**Abnormal Liver Function**

- **K70.*** (alcoholic liver disease)
- **K71.*** (toxic liver disease)
- **K72.*** (hepatic failure)
- **K73.*** (chronic hepatitis)
- **K74.*** (fibrosis and cirrhosis of liver)
- **K75.*** (other inflammatory liver diseases)
- **K76.*** (other diseases of liver)
- **K77** (liver disorders in diseases classified elsewhere)

**History of Bleeding**

- **D50-D64** (nutritional and hemolytic anemias)
  - D50.* (iron deficiency anemia)
  - D51.* (vitamin B12 deficiency anemia)
  - D52.* (folate deficiency anemia)
  - D53.* (other nutritional anemias)
  - D55-D59 (hemolytic anemias)
  - D60-D64 (aplastic and other anemias)
- **D62** (acute posthemorrhagic anemia)
- **D68.*** (other coagulation defects)
- **K20-K31** (diseases of esophagus, stomach and duodenum with bleeding)
  - K20.* (esophagitis)
  - K21.0 (gastro-esophageal reflux disease with esophagitis)
  - K22.6, K22.8 (gastro-esophageal laceration-hemorrhage syndrome)
  - K25-K28 (peptic ulcer disease - all sites)
  - K29.01, K29.21, K29.31, K29.41, K29.51, K29.61, K29.71, K29.81, K29.91 (gastritis with bleeding)
- **K31.811, K31.82** (angiodysplasia with bleeding)
- **K52.81** (eosinophilic colitis with bleeding)
- **K55.21** (angiodysplasia of colon with hemorrhage)
- **K57.01, K57.11, K57.13, K57.21, K57.31, K57.33, K57.41, K57.51, K57.53, K57.81, K57.91, K57.93** (diverticular disease with bleeding)
- **K62.5** (hemorrhage of anus and rectum)
- **K64.*** (hemorrhoids and perianal venous thrombosis)
- **K66.1** (hemoperitoneum)
- **K92.0, K92.1, K92.2** (gastrointestinal hemorrhage)
- **M25.0*** (hemarthrosis)
- **N02.*, N89.8, N92.0, N92.1, N93.8, N93.9, N95.0** (genitourinary bleeding)
- *R04.0, R04.1, R04.2, R04.8, R04.9** (hemorrhage from respiratory passages)
- **R31.*, R58** (hematuria, hemorrhage not elsewhere classified)
- *S06.4-S06.6*** (traumatic intracranial hemorrhage)

**Labile INR**

- **Z79.01** (long term use of anticoagulants)
- Clinical determination based on INR values, not specific ICD codes

**Alcohol Use**

- **F10.1*** (alcohol abuse)
- **F10.2*** (alcohol dependence)
- **F10.9*** (alcohol use, unspecified)

**eTable 5. Performance Metrics of Retrieval-Augmented Generation (RAG) versus Structured Data Approaches for Risk Factor Identification in Yale New Haven Health System (YNHHS).** Comprehensive evaluation of accuracy, precision, sensitivity, F1-score, specificity, and negative predictive value for all risk components.

| Condition | Approach | Accuracy (95% CI) | Precision (95% CI) | Sensitivity (95% CI) | F1-Score (95% CI) | Specificity (95% CI) | NPV (95% CI) |
| --- | --- | --- | --- | --- | --- | --- | --- |
| Hypertension | **RAG** | 0.952 (0.892-0.979) | 0.978 (0.925-0.994) | 0.968 (0.910-0.989) | 0.973 (0.922-0.991) | 0.800 (0.490-0.943) | 0.727 (0.434-0.903) |
|  | **Structured** | 0.856 (0.776-0.911) | 1.000 (0.954-1.000) | 0.840 (0.753-0.901) | 0.913 (0.843-0.954) | 1.000 (0.722-1.000) | 0.400 (0.234-0.593) |
| CHF | **RAG** | 0.942 (0.880-0.973) | 1.000 (0.912-1.000) | 0.870 (0.743-0.939) | 0.930 (0.864-0.965) | 1.000 (0.938-1.000) | 0.906 (0.810-0.956) |
|  | **Structured** | 0.875 (0.798-0.925) | 1.000 (0.896-1.000) | 0.717 (0.575-0.827) | 0.835 (0.752-0.894) | 1.000 (0.938-1.000) | 0.817 (0.712-0.890) |
| Diabetes | **RAG** | 0.990 (0.948-0.998) | 1.000 (0.912-1.000) | 0.976 (0.874-0.996) | 0.988 (0.943-0.997) | 1.000 (0.943-1.000) | 0.984 (0.917-0.997) |
|  | **Structured** | 0.923 (0.856-0.961) | 1.000 (0.896-1.000) | 0.805 (0.660-0.898) | 0.892 (0.818-0.938) | 1.000 (0.943-1.000) | 0.887 (0.793-0.942) |
| Stroke | **RAG** | 0.981 (0.933-0.995) | 1.000 (0.839-1.000) | 0.909 (0.722-0.975) | 0.952 (0.893-0.980) | 1.000 (0.955-1.000) | 0.976 (0.917-0.993) |
|  | **Structured** | 0.894 (0.820-0.940) | 1.000 (0.741-1.000) | 0.500 (0.307-0.693) | 0.667 (0.572-0.750) | 1.000 (0.955-1.000) | 0.882 (0.800-0.933) |
| Vascular | **RAG** | 0.981 (0.933-0.995) | 0.979 (0.889-0.996) | 0.979 (0.889-0.996) | 0.979 (0.930-0.994) | 0.982 (0.907-0.997) | 0.982 (0.907-0.997) |
|  | **Structured** | 0.885 (0.809-0.933) | 0.907 (0.784-0.963) | 0.830 (0.699-0.911) | 0.867 (0.788-0.919) | 0.930 (0.833-0.972) | 0.869 (0.762-0.932) |
| Uncontrolled HTN | **RAG** | 0.940 (0.875-0.972) | 0.783 (0.581-0.903) | 0.947 (0.754-0.991) | 0.857 (0.775-0.913) | 0.938 (0.864-0.973) | 0.987 (0.930-0.998) |
|  | **Structured** | 0.657 (0.562-0.743) | 0.818 (0.639-0.922) | 0.360 (0.240-0.500) | 0.500 (0.403-0.597) | 0.927 (0.831-0.973) | 0.614 (0.504-0.715) |
| Renal Disease | **RAG** | 0.990 (0.948-0.998) | 1.000 (0.867-1.000) | 0.962 (0.811-0.993) | 0.980 (0.932-0.995) | 1.000 (0.953-1.000) | 0.987 (0.932-0.998) |
|  | **Structured** | 0.750 (0.659-0.823) | 0.500 (0.332-0.668) | 0.577 (0.389-0.745) | 0.536 (0.440-0.629) | 0.808 (0.707-0.880) | 0.851 (0.753-0.915) |
| Liver Disease | **RAG** | 0.962 (0.905-0.985) | 0.921 (0.792-0.973) | 0.972 (0.858-0.995) | 0.946 (0.885-0.976) | 0.956 (0.878-0.985) | 0.985 (0.919-0.997) |
|  | **Structured** | 0.673 (0.578-0.756) | 0.750 (0.301-0.954) | 0.083 (0.029-0.218) | 0.150 (0.094-0.231) | 0.985 (0.921-0.997) | 0.670 (0.573-0.754) |
| Labile INR | **RAG** | 1.000 (1.000-1.000) | 1.000 (1.000-1.000) | 1.000 (1.000-1.000) | 1.000 (1.000-1.000) | 1.000 (1.000-1.000) | 1.000 (1.000-1.000) |
|  | **Structured** | 0.837 (0.754-0.895) | 0.077 (0.014-0.333) | 0.167 (0.030-0.564) | 0.105 (0.060-0.179) | 0.878 (0.798-0.929) | 0.945 (0.878-0.976) |
| Bleeding History | **RAG** | 0.971 (0.919-0.990) | 1.000 (0.845-1.000) | 0.875 (0.690-0.957) | 0.933 (0.868-0.967) | 1.000 (0.954-1.000) | 0.964 (0.899-0.988) |
|  | **Structured** | 0.740 (0.649-0.815) | 0.435 (0.256-0.632) | 0.417 (0.245-0.612) | 0.426 (0.335-0.522) | 0.838 (0.742-0.903) | 0.827 (0.731-0.894) |
| Bleeding Meds | **RAG** | 0.962 (0.905-0.985) | 0.985 (0.921-0.997) | 0.957 (0.881-0.985) | 0.971 (0.918-0.990) | 0.971 (0.851-0.995) | 0.917 (0.782-0.971) |
|  | **Structured** | 0.683 (0.588-0.764) | 0.687 (0.590-0.770) | 0.971 (0.902-0.992) | 0.805 (0.718-0.869) | 0.088 (0.030-0.230) | 0.600 (0.231-0.882) |
| Alcohol Use Disorder | **RAG** | 0.990 (0.948-0.998) | 1.000 (0.757-1.000) | 0.923 (0.667-0.986) | 0.960 (0.903-0.984) | 1.000 (0.959-1.000) | 0.989 (0.941-0.998) |
|  | **Structured** | 0.885 (0.809-0.933) | 1.000 (0.207-1.000) | 0.077 (0.014-0.333) | 0.143 (0.088-0.223) | 1.000 (0.959-1.000) | 0.883 (0.807-0.932) |

**eTable 6. Performance Metrics of Retrieval-Augmented Generation (RAG) versus Structured Data Approaches for Risk Factor Identification in Medical Information Mart for Intensive Care (MIMIC-IV).** Comprehensive evaluation of accuracy, precision, sensitivity, F1-score, specificity, and negative predictive value for all risk components.

| Condition | Approach | Accuracy (95% CI) | Precision (95% CI) | Sensitivity (95% CI) | F1-Score (95% CI) | Specificity (95% CI) | NPV (95% CI) |
| --- | --- | --- | --- | --- | --- | --- | --- |
| Hypertension | **RAG** | 0.940 (0.875-0.972) | 0.953 (0.886-0.982) | 0.976 (0.917-0.993) | 0.965 (0.908-0.987) | 0.750 (0.505-0.898) | 0.857 (0.601-0.960) |
|  | **Structured** | 0.920 (0.850-0.959) | 0.953 (0.886-0.982) | 0.953 (0.886-0.982) | 0.953 (0.893-0.981) | 0.714 (0.454-0.883) | 0.714 (0.454-0.883) |
| CHF | **RAG** | 0.960 (0.902-0.984) | 0.925 (0.821-0.970) | 1.000 (0.927-1.000) | 0.961 (0.903-0.985) | 0.922 (0.815-0.969) | 1.000 (0.924-1.000) |
|  | **Structured** | 0.890 (0.814-0.937) | 0.887 (0.774-0.947) | 0.904 (0.794-0.958) | 0.895 (0.820-0.941) | 0.875 (0.753-0.941) | 0.894 (0.774-0.954) |
| Diabetes | **RAG** | 0.970 (0.915-0.990) | 0.962 (0.870-0.989) | 0.980 (0.897-0.997) | 0.971 (0.917-0.990) | 0.959 (0.863-0.989) | 0.979 (0.891-0.996) |
|  | **Structured** | 0.740 (0.646-0.816) | 0.500 (0.369-0.631) | 1.000 (0.871-1.000) | 0.667 (0.570-0.751) | 0.649 (0.535-0.748) | 1.000 (0.926-1.000) |
| Stroke/TIA | **RAG** | 0.930 (0.863-0.966) | 0.862 (0.694-0.945) | 0.893 (0.728-0.963) | 0.877 (0.799-0.928) | 0.944 (0.866-0.978) | 0.958 (0.883-0.986) |
|  | **Structured** | 0.840 (0.756-0.899) | 0.552 (0.375-0.716) | 0.842 (0.624-0.945) | 0.667 (0.570-0.751) | 0.840 (0.745-0.904) | 0.958 (0.883-0.986) |
| Vascular Disease | **RAG** | 0.940 (0.875-0.972) | 0.970 (0.896-0.992) | 0.941 (0.858-0.977) | 0.955 (0.895-0.982) | 0.938 (0.799-0.983) | 0.882 (0.734-0.953) |
|  | **Structured** | 0.510 (0.413-0.606) | 0.258 (0.167-0.374) | 1.000 (0.816-1.000) | 0.410 (0.318-0.508) | 0.410 (0.310-0.517) | 1.000 (0.898-1.000) |
| Uncontrolled HTN | **RAG** | 0.940 (0.875-0.972) | 0.783 (0.581-0.903) | 0.947 (0.754-0.991) | 0.857 (0.775-0.913) | 0.857 (0.775-0.913) | 0.938 (0.864-0.973) |
|  | **Structured** | 0.660 (0.563-0.745) | 0.870 (0.679-0.955) | 0.392 (0.270-0.529) | 0.541 (0.443-0.635) | 0.939 (0.835-0.979) | 0.597 (0.486-0.700) |
| Renal Disease | **RAG** | 0.960 (0.902-0.984) | 0.938 (0.799-0.983) | 0.938 (0.799-0.983) | 0.938 (0.872-0.971) | 0.971 (0.899-0.992) | 0.971 (0.899-0.992) |
|  | **Structured** | 0.790 (0.700-0.858) | 0.594 (0.423-0.745) | 0.704 (0.515-0.841) | 0.644 (0.546-0.731) | 0.822 (0.719-0.893) | 0.882 (0.785-0.939) |
| Liver Disease | **RAG** | 0.990 (0.946-0.998) | 0.933 (0.702-0.988) | 1.000 (0.785-1.000) | 0.966 (0.909-0.987) | 0.988 (0.937-0.998) | 1.000 (0.957-1.000) |
|  | **Structured** | 0.560 (0.462-0.653) | 1.000 (0.706-1.000) | 0.254 (0.161-0.378) | 0.405 (0.314-0.503) | 1.000 (0.914-1.000) | 0.482 (0.379-0.587) |
| Labile INR | **RAG** | 0.990 (0.946-0.998) | 0.933 (0.717-0.989) | 1.000 (0.796-1.000) | 0.968 (0.912-0.989) | 0.988 (0.936-0.998) | 1.000 (0.956-1.000) |
|  | **Structured** | 0.440 (0.347-0.538) | 1.000 (0.806-1.000) | 0.222 (0.142-0.331) | 0.364 (0.276-0.461) | 1.000 (0.879-1.000) | 0.333 (0.242-0.439) |
| Bleeding History | **RAG** | 0.890 (0.814-0.937) | 0.853 (0.750-0.918) | 0.983 (0.910-0.997) | 0.913 (0.842-0.954) | 0.756 (0.607-0.862) | 0.969 (0.843-0.994) |
|  | **Structured** | 0.580 (0.482-0.672) | 0.515 (0.398-0.629) | 0.795 (0.655-0.888) | 0.625 (0.527-0.714) | 0.411 (0.292-0.541) | 0.719 (0.546-0.844) |
| Bleeding Meds | **RAG** | 0.970 (0.915-0.990) | 0.967 (0.908-0.989) | 1.000 (0.959-1.000) | 0.983 (0.935-0.996) | 0.727 (0.434-0.903) | 1.000 (0.676-1.000) |
|  | **Structured** | 0.910 (0.838-0.952) | 0.989 (0.941-0.998) | 0.919 (0.849-0.958) | 0.953 (0.892-0.980) | 0.000 (0.000-0.793) | 0.000 (0.000-0.324) |
| Alcohol Use Disorder | **RAG** | 1.000 (0.963-1.000) | 1.000 (0.566-1.000) | 1.000 (0.566-1.000) | 1.000 (0.963-1.000) | 1.000 (0.961-1.000) | 1.000 (0.961-1.000) |
|  | **Structured** | 0.970 (0.915-0.990) | 1.000 (0.566-1.000) | 0.625 (0.306-0.863) | 0.769 (0.678-0.841) | 1.000 (0.960-1.000) | 0.968 (0.911-0.989) |

**eTable 7: Comparison of Processing Times for Manual Annotation versus Retrieval-Augmented Generation (RAG) Approach**

| **Metric** | **Manual Annotation** | **RAG-LLM Approach** |
| --- | --- | --- |
| **Mean time per patient** | 12 minutes (range 10-15 minutes) | 35 seconds (range: 28-45 seconds) |
| **Validation cohort (n=300)** | 60 person-hours | 2.9 computational hours |
| **Computational resources** | - | Server with 8x NVIDIA H100 GPU, 80GB RAM, 96 CPU cores |
| **Parallel processing** | - | 8 parallel threads |

**eTable 8. Concordance and Discordance Patterns Between Manual Clinician Review and Retrieval-Augmented Generation (RAG) Assessment for CHA₂DS₂-VASc and HAS-BLED Risk Factors in Validation Cohorts.**

| **Characteristic** | **YNHHS Cohort (n=200)** | **MIMIC-IV Cohort (n=100)** |
| --- | --- | --- |
| **Total assessments** | 2,400 | 1,200 |
| **Concordant assessments** | 2,196 (91.5%) | 1,073 (89.4%) |
| **Discordant assessments** | 204 (8.5%) | 127 (10.6%) |
| **RAG correct (manual error)** | 78 (38.2% of discordance) | 42 (33.1% of discordance) |
| **Manual correct (RAG error)** | 126 (61.8% of discordance) | 85 (66.9% of discordance) |

**eTable 9. MIMIC Error Pattern Characterization in Retrieval-Augmented Generation (RAG) and Manual Clinical Review with Representative Clinical Examples.**

| **Error Category** | **Error Pattern** | **YNHHS Frequency n (%)** | **MIMIC Frequency n (%)** | **Clinical Example** |
| --- | --- | --- | --- | --- |
| **Manual Validation Errors** | Laboratory values outside of thresholds | 32 (41.0%) | 17 (40.5%) | Creatinine 2.3 mg/dL exceeding 1.5 mg/dL threshold for renal dysfunction |
|  | Subspecialty documentation review limitations | 22 (28.2%) | 12 (28.6%) | Explicit mention of "chronic systolic heart failure" in cardiology consultation note |
|  | Complex pathology classification | 15 (19.2%) | 8 (19.0%) | Stable ascending aortic aneurysm (4.5 cm) not classified as vascular disease |
|  | Medication regimen interpretation | 9 (11.5%) | 5 (11.9%) | Multiple INR values outside therapeutic range (2.0-3.0) without recognizing pattern of labile anticoagulation |
|  | **Total Manual Errors** | **78** | **42** |  |
| **RAG Assessment Errors** | Contextual interpretation of isolated findings | 35 (27.8%) | 24 (28.2%) | Isolated systolic BP reading of 155 mmHg incorrectly classified as uncontrolled hypertension |
|  | Temporal disambiguation challenges | 28 (22.2%) | 19 (22.4%) | Discontinued antiplatelet therapy incorrectly flagged as current bleeding medication |
|  | Acute vs. chronic condition differentiation | 24 (19.0%) | 16 (18.8%) | Acute kidney injury misclassified as chronic renal dysfunction |
|  | Specificity in bleeding risk assessment | 21 (16.7%) | 14 (16.5%) | Prophylactic anticoagulation during hospitalization misclassified as therapeutic anticoagulation |
|  | Documentation inconsistency resolution | 18 (14.3%) | 12 (14.1%) | Contradictory documentation of liver function across multiple notes |
|  | **Total RAG Errors** | **126** | **85** |  |

**eTable 10 Differential Risk Factor Identification Between Retrieval-Augmented Generation (RAG) and Structured Data Approaches in the Validation Subset.**

| **Component** | **YNHHS Structured Count** | **YNHHS RAG Count** | **YNHHS Ground Truth Count** | **YNHHS Difference** | **MIMIC-IV Structured Count** | **MIMIC-IV RAG Count** | **MIMIC-IV Ground Truth Count** | **MIMIC-IV Difference** |
| --- | --- | --- | --- | --- | --- | --- | --- | --- |
| **CHA₂DS₂-VASc Components** |  |  |  |  |  |  |  |  |
| **Congestive Heart Failure** | 31 | 46 | 46 | +15 | 56 | 56 | 53 | 0 |
| **Hypertension** | 79 | 82 | 87 | +3 | 88 | 97 | 86 | +9 |
| **Diabetes** | 34 | 37 | 39 | +3 | 33 | 62 | 52 | +29 |
| **Stroke/TIA** | 10 | 27 | 26 | +17 | 32 | 43 | 29 | +11 |
| **Vascular Disease** | 42 | 54 | 54 | +12 | 61 | 72 | 66 | +11 |
| **HAS-BLED Components** |  |  |  |  |  |  |  |  |
| **Uncontrolled Hypertension** | 7 | 7 | 7 | 0 | 19 | 19 | 19 | 0 |
| **Renal Disease** | 28 | 30 | 29 | +2 | 32 | 35 | 32 | +3 |
| **Liver Disease** | 6 | 15 | 15 | +9 | 36 | 41 | 26 | +5 |
| **Bleeding History** | 21 | 58 | 56 | +37 | 49 | 75 | 68 | +26 |
| **Medication Use** | 87 | 85 | 81 | -2 | 93 | 99 | 91 | +6 |
| **Alcohol Use** | 2 | 9 | 9 | +7 | 0 | 11 | 11 | +11 |

**eTable 11. Risk Score Reclassification Analysis Following RAG-LLM Implementation in the YNHHS Cohort**

| **Original Risk Category (Structured Data)** | **New Risk Category (RAG-LLM)** | | | **Total** | **Reclassification Rate** |
| --- | --- | --- | --- | --- | --- |
| **CHA₂DS₂-VASc** |  |  |  |  |  |
|  | **Low** | **Intermediate** | **High** |  |  |
| **Low** | 93 (31.3 %) | 90 (30.3 %) | 114 (38.4 %) | 297 | 204 / 297 (68.7 %) |
| **Intermediate** | 23 (0.8 %) | 1,008 (34.6 %) | 1,879 (64.6 %) | 2,910 | 1,902 / 2,910 (65.4 %) |
| **High** | 0 (0.0 %) | 184 (0.2 %) | 100,813 (99.8 %) | 100,997 | 184 / 100,997 (0.2 %) |
| **Total** | **116** | **1,282** | **102,806** | **104,204** | **2,290 / 104,204 (2.2 %)** |
| **HAS-BLED** |  |  |  |  |  |
|  | **Low** | **Intermediate** | **High** |  |  |
| **Low** | 34,067 (67.0%) | 13,901 (27.4%) | 2,806 (5.5%) | 50,774 | 16,707 / 50,774 (33.0%) |
| **Intermediate** | 4,741 (14.6%) | 15,901 (49.0%) | 11,764 (36.3%) | 32,406 | 16,505 / 32,406 (50.9%) |
| **High** | 0 (0.0%) | 3,654 (17.3%) | 17,370 (82.7%) | 21,024 | 3,654 / 21,024 (17.4%) |
| **Total** | **38,739** | **33,456** | **31,940** | **104,204** | **36,866 / 104,204 (35.4%)** |

**eTable 12. Risk Score Reclassification Analysis Following RAG-LLM Implementation in the MIMIC-IV Cohort**

| **Original Risk Category (Structured Data)** | **New Risk Category (RAG-LLM)** | | | **Total** | **Reclassification Rate** |
| --- | --- | --- | --- | --- | --- |
| **CHA₂DS₂-VASc** |  |  |  |  |  |
|  | **Low** | **Intermediate** | **High** |  |  |
| **Low** | 573 (49.5 %) | 347 (30.0 %) | 238 (20.6 %) | 1,158 | 585 / 1,158 (50.5 %) |
| **Intermediate** | 9 (0.3 %) | 1,135 (33.9 %) | 2,206 (65.9 %) | 3,350 | 2,215 / 3,350 (66.1 %) |
| **High** | 7 (0.1 %) | 63 (0.7 %) | 8,539 (99.2 %) | 8,609 | 70 / 8,609 (0.8 %) |
| **Total** | **589** | **1,545** | **10,983** | **13,117** | **2,870 / 13,117 (21.9 %)** |
| **HAS-BLED** |  |  |  |  |  |
|  | **Low** | **Intermediate** | **High** |  |  |
| **Low** | 3,702 (44.4%) | 2,996 (35.9%) | 1,639 (19.7%) | 8,337 | 4,635 / 8,337 (55.6%) |
| **Intermediate** | 149 (4.4%) | 1,388 (40.7%) | 1,874 (54.9%) | 3,411 | 2,023 / 3,411 (59.3%) |
| **High** | 6 (0.4%) | 40 (2.9%) | 1,323 (96.7%) | 1,369 | 46 / 1,369 (3.4%) |
| **Total** | **4,855** | **4,748** | **3,514** | **13,117** | **6,704 / 13,117 (51.1%)** |

**eTable 13. Risk Score Reclassification Analysis Following RAG-LLM Implementation in the YNHHS No Anticoagulation Subset**

|  | **RAG Low** | **RAG Intermediate** | **RAG High (≥3)** | **Row Total** | **Row % High** |
| --- | --- | --- | --- | --- | --- |
| **CHA₂DS₂-VASc** |  |  |  |  |  |
| Structured Low (n=297) | 93 (31.3%) | 90 (30.3%) | 114 (38.4%) | 297 | 38.4% |
| Structured Intermediate (n=2,910) | 22 (0.8%) | 992 (34.1%) | 1,896 (65.1%) | 2,910 | 65.1% |
| Structured High (n=100,997) | 0 (0.0%) | 174 (0.2%) | 100,823 (99.8%) | 100,997 | 99.8% |
| **HAS-BLED** |  |  |  |  |  |
| Structured Low (n=16,613) | 7,064 (42.5%) | 5,401 (32.5%) | 4,148 (24.9%) | 16,613 | 24.9% |
| Structured Intermediate (n=32,251) | 12,777 (39.6%) | 10,325 (32.0%) | 9,149 (28.4%) | 32,251 | 28.4% |
| Structured High (n=55,340) | 18,967 (34.3%) | 17,730 (32.0%) | 18,643 (33.7%) | 55,340 | 33.7% |
| **Newly High CHA₂DS₂-VASc (n=2,010)** | 732 (36.4%) | 684 (34.0%) | 594 (29.6%) | 2,010 | 29.6% |
| **Structured Antithrombotic Use (n=18,512)** | 2,062 (11.1%) | 5,786 (31.3%) | 10,664 (57.6%) | 18,512 | 57.6% |

**eFigure 1. Native EHR Schemas (YNHHS and MIMIC-IV) and Automated Schema Mapping (Table and Column Mapping)**

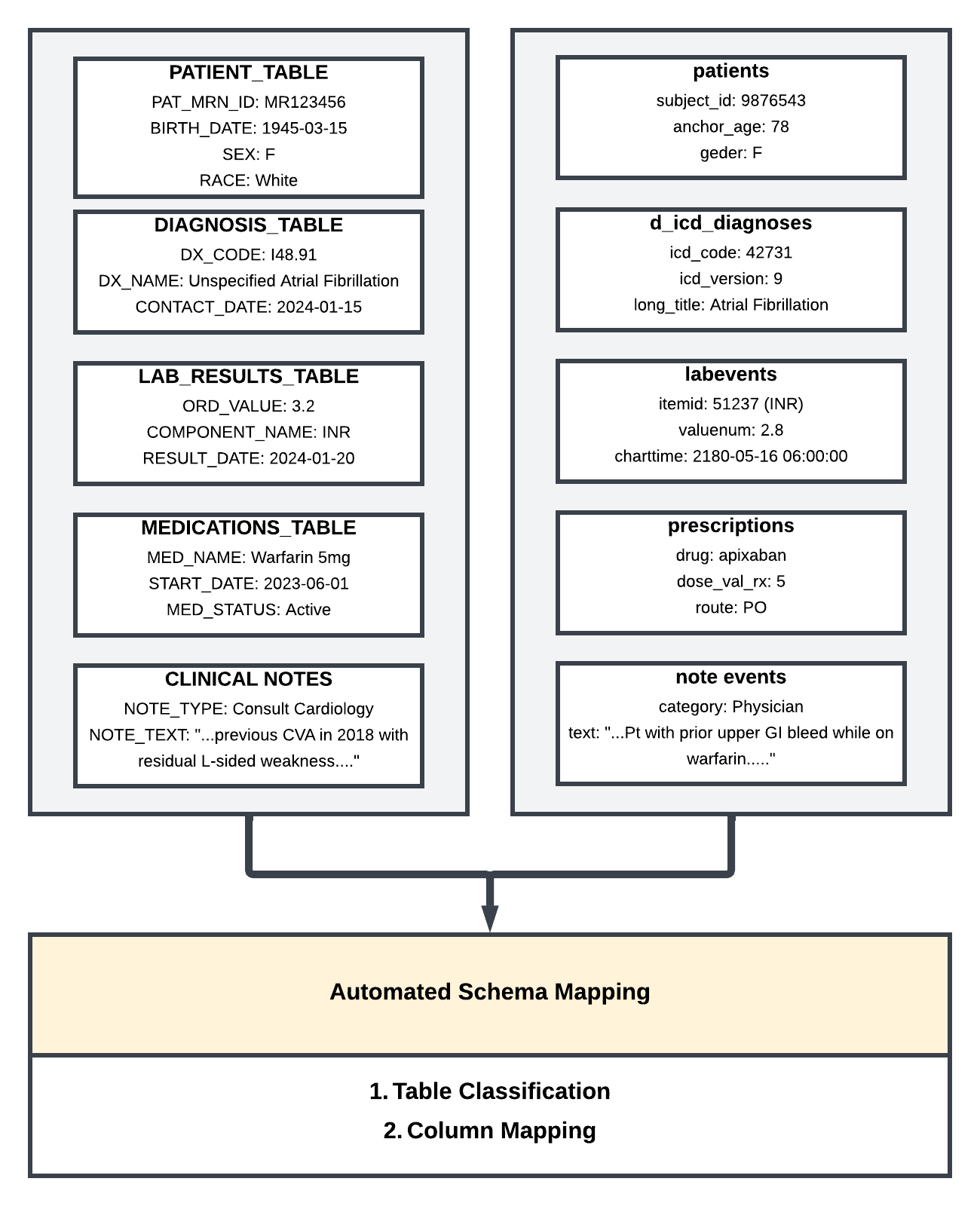

**eFigure 2. Query Template Construction Methodology for Retrieval-Augmented Generation (RAG).**
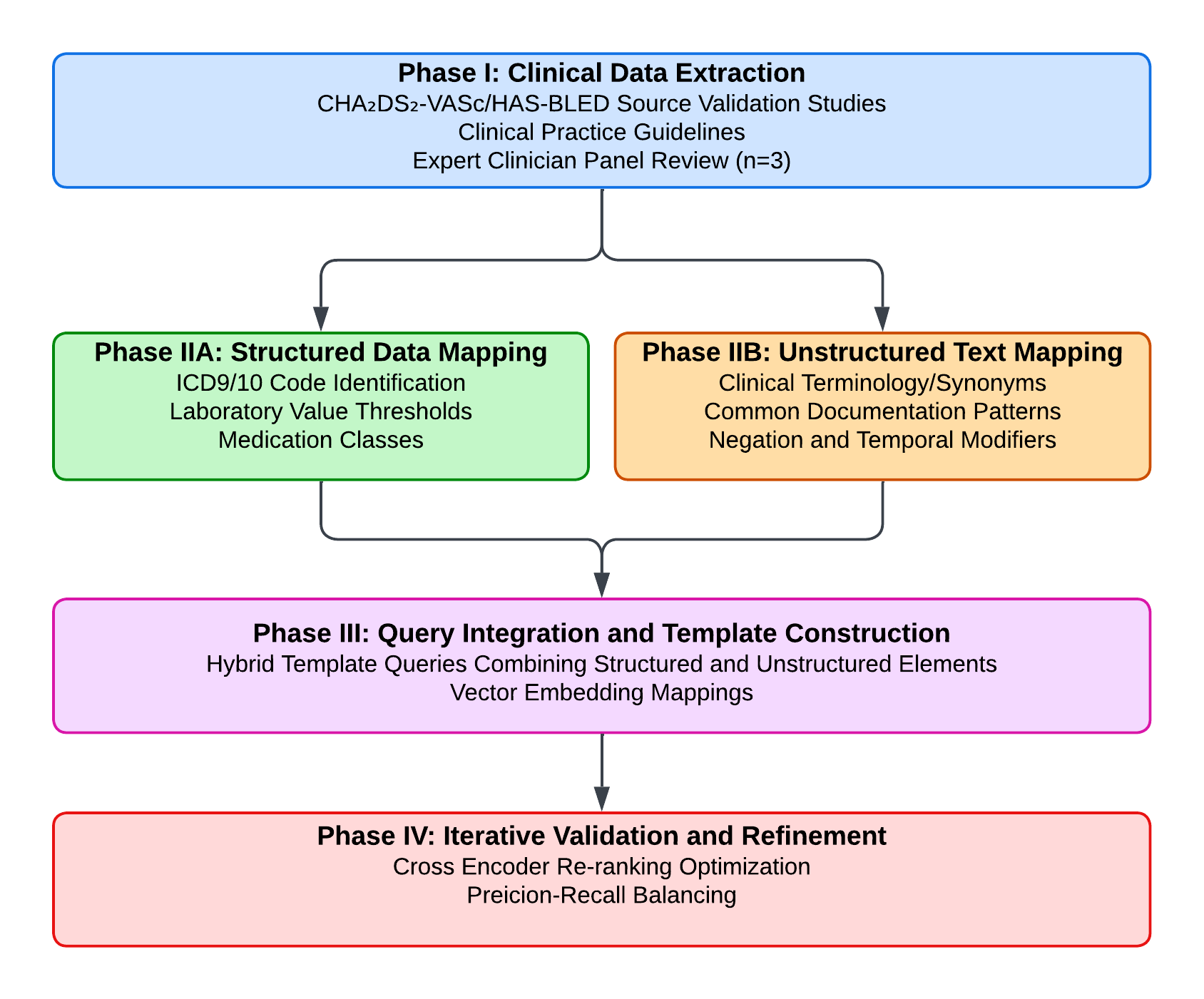

**eFigure 3.** **Clinical Text Processing Pipeline for Retrieval-Augmented Generation (RAG) Implementation Showing Document Indexing and Query Augmentation Process**

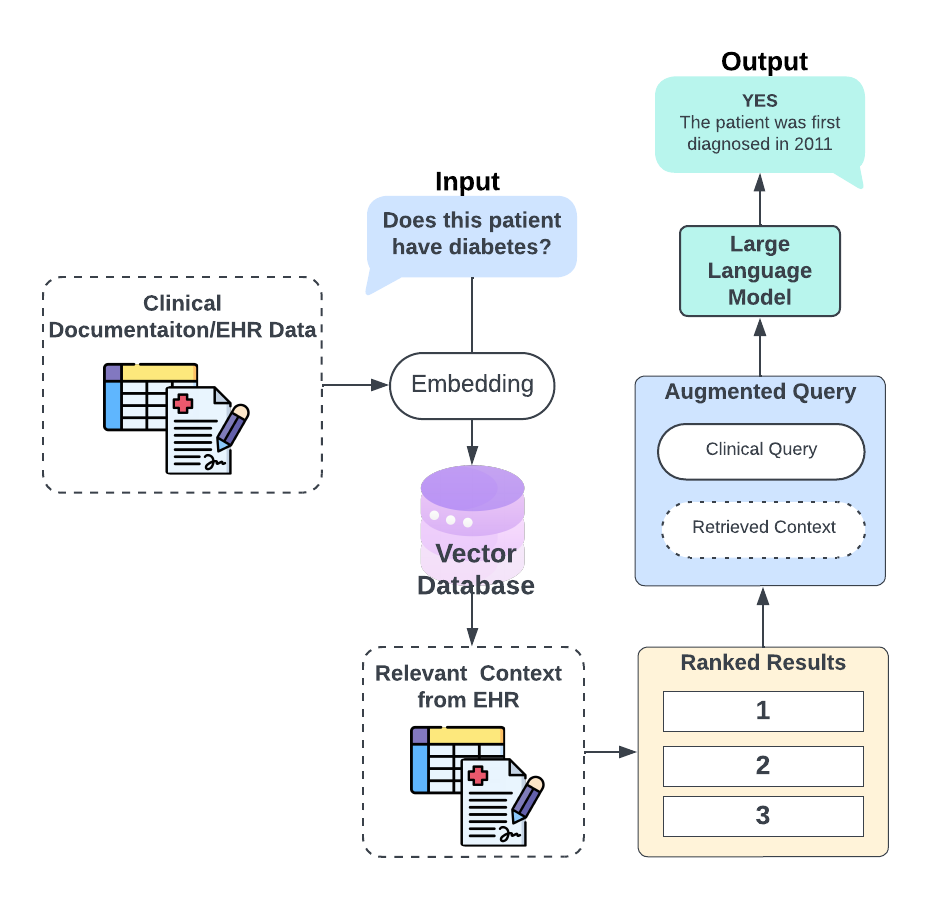

**eFigure 4: Risk Score Changes with RAG-Enhanced Detection for MIMIC-IV Patients with Atrial Fibrillation (n=13,117)**

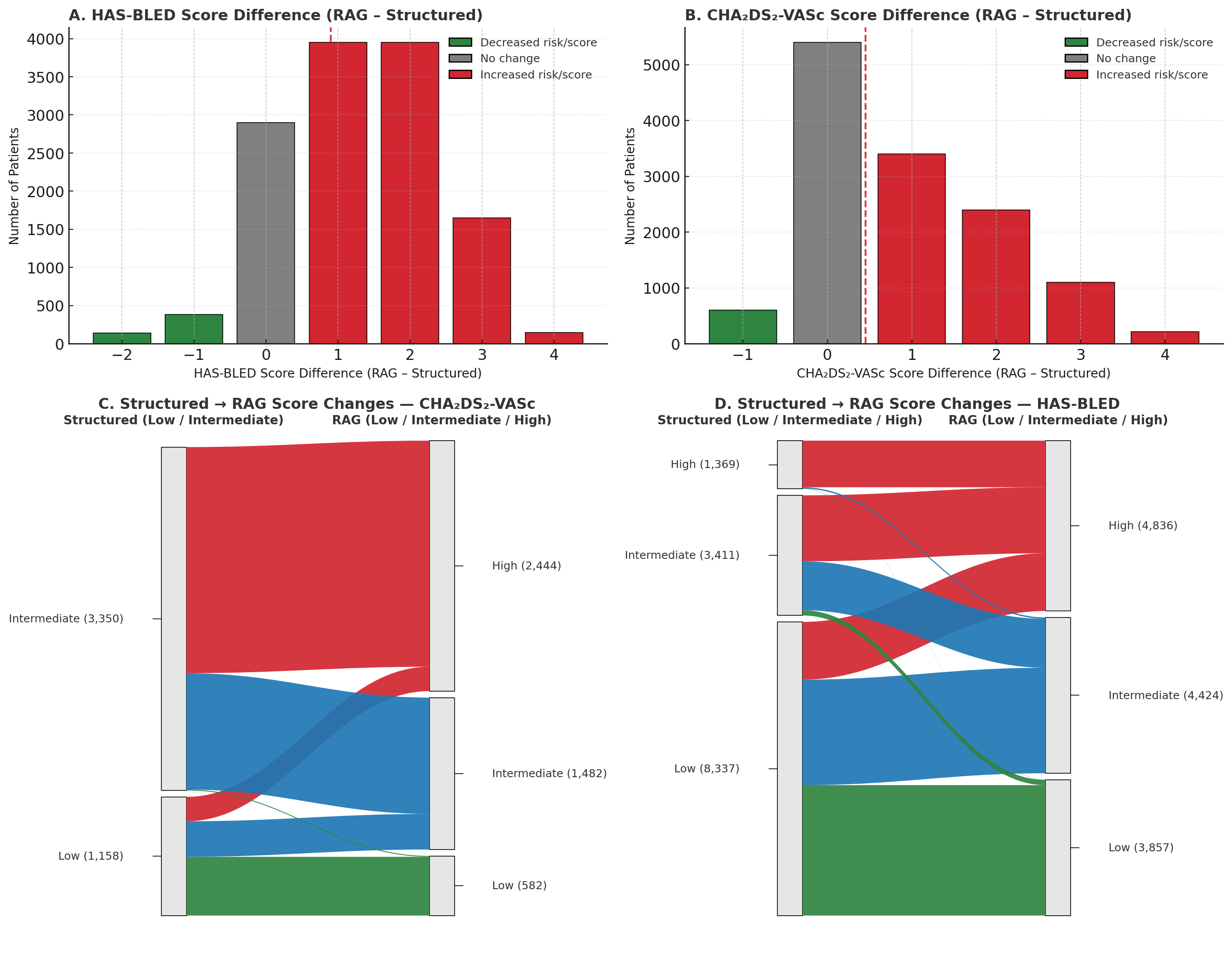
